# Modelling drug resistance emergence and transmission in HIV-1 in the UK

**DOI:** 10.1101/2023.04.24.23288580

**Authors:** Anna Zhukova, David Dunn, Olivier Gascuel, the UK HIV Drug Resistance Database & the Collaborative HIV, Anti-HIV Drug Resistance Network

## Abstract

A deeper understanding of HIV-1 transmission and drug resistance mechanisms can lead to improvement in current treatment policies. However, the rates at which HIV-1 drug resistance mutations (DRMs) are acquired and at which transmitted DRMs persist are multi-factorial and vary considerably between different mutations. We develop a method for estimation of drug resistance acquisition and transmission patterns, which refines the method we described in Mourad et al. AIDS 2015. The method uses maximum likelihood ancestral character reconstruction informed by treatment roll-out dates and allows for analysis of very large data sets. We apply our method to transmission trees reconstructed on the data obtained from the UK HIV drug resistance database to make predictions for known DRMs. Our results show important differences between DRMs, in particular between polymorphic and non-polymorphic DRMs, and between the B and C subtypes. Our estimates of reversion times, based on a very large number of sequences, are compatible but more accurate than those already available in the litterature, with narrower confidence intervals. We consistently find that large resistance clusters are associated with polymorphic DRMs and DRMs with long loss time, which require special surveillance. As in other high-income countries (e.g. Switzerland), the prevalence of sequences with DRMs is decreasing, but among these, the fraction of transmitted resistance is clearly increasing compared to the fraction of acquired resistance mutations. All this indicates that efforts to monitor these mutations and the emergence of resistance clusters in the population must be maintained in the long term.

## 1. Introduction

Drug resistance is an increasing health problem. *Drug resistance mutations (DRMs)* emerge in HIV viruses through selective pressure during *antiretroviral therapy (ART)* and make the current ART drug combination ineffective both for sustaining the patient’s well-being and for prevention of virus transmission [1,2]. Drug resistant viruses can therefore be transmitted to treatment-naive patients, who in turn can transmit them further [3,4], endangering the efficacy of treatment for the whole population. The rates at which DRMs are acquired and *transmitted drug resistance (TDR)* mutations persist are likely to be multi-factorial and have been shown to vary considerably depending on duration and type of treatment, and mutations [5]. Hence, having a deeper understanding of HIV transmission and drug resistance mechanisms is important as it can lead to improvement in current treatment policies [6].

Phylodynamics uses phylogenetic trees (i.e. genealogies of the pathogen population) inferred from the pathogen sequence data to estimate the epidemiological parameters. Several phylodynamic models of pathogen transmission were developed [7–10].

An important trade-off in phylodynamic modeling is between the complexity of the biological questions that a model can address and its computational speed. On one side of the spectrum there are computationally-light statistical approaches, such as the study by Mourad *et al*. [4] of persistence times of drug-resistance in the HIV-1-infected untreated population in the UK. The analysis used a a parsimony-based approach [11] to extract “phylotypes” of sequences, the most recent common ancestor of which was bearing a resistant mutation that is still shared by the majority of the sequences in the phylotype. Once dated and combined with the treatment-naive/experienced status, these phylotypes were used to zoom on the most readable parts of the phylogeny and compute simple statistics which are immediately accessible from the annotated tree. The simplicity of the method makes it computationally very efficient. It was applied to a large set of ≈ 25, 000 HIV-1 subtype B sequences from the UK, where it showed that around 70% of transmitted drug-resistance had a treatment-naive source.

However to address more refined questions such as estimation of rates of different events (transmission, drug resistance acquisition, etc.), more complex methods are needed, such as modeling of the viral dynamics with ordinary differential equations (ODEs). Kühnert *et al*. [9] proposed a piecewise-constant two-type (resistant and sensitive) birth–death model to estimate the fitness cost of DRMs. The fitness was measured as a ratio between transmission rates of hosts infected by drug resistant strains and transmission rates of hosts infected by sensitive strains. They applied this model to the data from the Swiss HIV cohort study. They reconstructed a maximum likelihood tree for 5 638 *pol*-gene sequences from the Swiss HIV cohort study and 4 284 closely related sequences from the Los Alamos HIV database. On this tree, for each of 15 major DRMs present in the Swiss cohort sequences, they identified its transmission clusters of up to 250 sequences each, containing > 80% of Swiss sequences and at least one sequence with the mutation. Kühnert *et al*. [9] estimated the model parameters on all the clusters for each mutation separately, in a Bayesian setting. To account for the fact that DRMs appear under aniretroviral (ARV) selective pressure, they put the rates of state change (from sensitive to resistant and vice versa) to zero before significant usage of the related drug(s) in Switzerland. The study showed that some of the mutations (RT:D67N, RT:K70R, RT:M184V, RT:K219Q) decreased the fitness, one (PR:L90M) seemed to increase the fitness, while the others did not have a significant effect.

The models above, and more generally the family of multi-type birth-death models [7] with a Bayesian birth-death skyline plot (allowing the parameters to change in a piece-wise constant manner) [12] it belongs to, define ODEs for fine-tuned parameter estimation. However, their complexity prevents resolving them analytically. Numerical solution of the ODEs, on the other hand, takes long computational time and prevents the application of these models to larger datasets (dozens of thousands of sequences), while larger data sets are desirable for more accurate parameter estimation.

A compromise between the model complexity and computational speed when applied to large datasets needs to be found. In this study we propose such a compromise that improves the approach by Mourad *et al*. [4] by using maximum likelihood and combining it with the skyline ideas of Stadler *et al*. [8], for analysis of DRM transmission patterns.

Our approach uses *ancestral character reconstruction (ACR)* on a partially sampled transmission tree. Using ancestral scenario reconstruction tool PastML [13], we study ancestral states for presence/absence of common surveillance DRMs. In a tree annotated with PastML, we can discriminate between two types of resistant nodes: (1) those whose parent node does not have the DRM, which correspond to *acquired drug resistance (ADR)*, and (2) those whose parent node is also resistant, such nodes form TDR clusters. We also identify the scenarios of DRM loss (when the parent node has the mutation, while the child does not). Moreover, we account for the changes in treatment policies by allowing for separate ACR for different time intervals (e.g. before and after the first DRM-provoking ARV introduction). Once the reconstruction is performed we visualize the results with PastML and calculate various statistics for transmission patterns.

We apply our approach to analyze the patterns of DRM emergence, transmission and loss in HIV-1-infected individuals in the UK, using sequences and metadata from the UK HIV Drug Resistance Database [14].

## 2. Materials and Methods

The UK HIV Drug Resistance Database provides HIV protease (PR) and reverse transcriptase (RT) sequences extracted during the resistance tests and the corresponding metadata (e.g., treatment status of the patient before the test: treatment-experienced, -naive, or unknown; and date of the test).

In response to our request for data from the database we obtained 88 009 sequences for 60 846 different patients, sampled between 1996 and 2016.

### 2.0.1. Sequence subtyping and alignment

We subtyped (pure subtypes and recombination positions) and aligned the sequences against the Los Alamos 2010 subtype reference *pol*-gene alignment [15] using jpHMM [16] (for detailed options see Appendix A).

All together, we obtained a large alignment of 88 009 sequences, from which we extracted the alignments for the B and C subtypes. We filtered them to contain only the first sequence (in terms of sampling date) when several sequences were present for the same patient. We hence obtained a 40 055-sequence alignment for the B subtype, and a 19 139-sequence alignment for the C subtype. To each of them we added five randomly selected HIV-1 group M sequences of other pure subtypes to be used as an outgroup for tree rooting.

### 2.0.2. Transmission tree reconstruction

We reconstructed phylogenetic trees for B and C sequences separately, using RAxML-NG (v0.9.0, evolutionary model GTR+G4+FO+IO, for detailed options see Appendix A) [17] and rooted them with the outgroup sequences, which we then removed. For tree reconstruction, the positions of surveillance DRMs were removed from the alignment, as they are influenced by treatment-selection forces unlike the other positions, and could bias the reconstruction by grouping together the sequences that share the same DRMs.

We then dated each tree with LSD2 [18] (v2.3: github.com/tothuhien/lsd2/tree/v1.4.2.2, under strict molecular clock with outlier removal, for detailed options see Appendix A) using tip sampling dates.

### 2.0.3. Ancestral character reconstruction

For each DRM (surveillance or accessory) listed in the Stanford HIV Drug Resistance Database [19] we extracted its presence/absence in the sequences of our data sets and the ARVs that can provoke it with Sierra, the Stanford Algorithm [20] web service. We then analyzed the DRMs that were found in at least 0.5% of sequences (after filtering by patient and temporal outlier removal) of our dataset (either B or C, analyzed separately).

Each DRM name (e.g. RT:T215D) contain 2 pieces of information: the DRM position (e.g. RT:T215, which in turn contains the protein name: RT (reverse transcriptase) or PR (protease), the reference position of the amino acid, e.g. 215, and its wildtype amino acid, e.g. T), and its mutated amino acid, associated with resistance (e.g. D).

We analyzed each DRM position independently by reconstructing its states in the ancestral nodes, based on the tip states. The DRMs with prevalence > 0.5% found in this position in the dataset of interest (e.g. B) were analyzed together. For the majority of the DRM positions, only one DRM with prevalence > 0.5% was found (e.g. PR:L90M for the position PR:L90), however for the positions RT:T215 and RT:K219 in the B data set and for the position RT:V179 in the C data set, several DRMs were found (RT:T215D/F/S/Y, RT:K219E/N/Q, and RT:V179D/E).

Possible states for ancestral character reconstruction (ACR) corresponded to DRM presence (i.e. the resistant state) or absence (sensitive state) for DRM positions with only one DRM. For instance, for PR:L90M the resistant state corresponds to the amino acid M, and the sensitive state to any other amino acid; in practice, the sensitive state is almost uniquely L. In the B data set, 97.79% of sequences have L at the position PR:90; 1.93% have M; less than 0.01% have W or F, and 0.23% have an ambiguity at this position (so their initial state for ACR is unresolved between sensitive and resistant). For positions with several DRMs, the resistant state was split into all the possibilities (e.g., D, F, S or Y for RT:T215).

For polymorphic mutations (e.g. RT:S68G), ACR was performed on the corresponding (B or C) time-scaled tree with PastML (v1.9.40, MAP (maximum a posteriori) decision rule) without taking into account the year of ARV acceptance, as the these mutations could be present independently of ARVs.

To reconstruct the ancestral character states for non-polymorphic DRMs, we used the procedure visualised in Figure 1a (which we first proposed and applied to study HIV resistance patterns in Cuba in [21]). For each ARV we extracted the dates of their acceptance with Wikipedia python package (https://github.com/goldsmith/Wikipedia). We cut the time-scaled tree at the earliest of the dates of acceptance of ARVs that can provoke the DRM (e.g. for PR:L90M, saquinavir (SQV) was accepted in 1995). We hence obtained the pre-treatment-introduction tree and a forest of post-treatment-introduction subtrees. For the trees in the forest we added additional one-child root nodes (as parents of the corresponding tree roots, at distances that corresponded to the differences between the root dates and the ARV acceptance date), which we marked as sensitive in the PastML input annotation file. We performed ACR with PastML on the forest, and then combined it with the all-sensitive annotation for the pre-treatment-introduction tree nodes.

**Figure 1.**
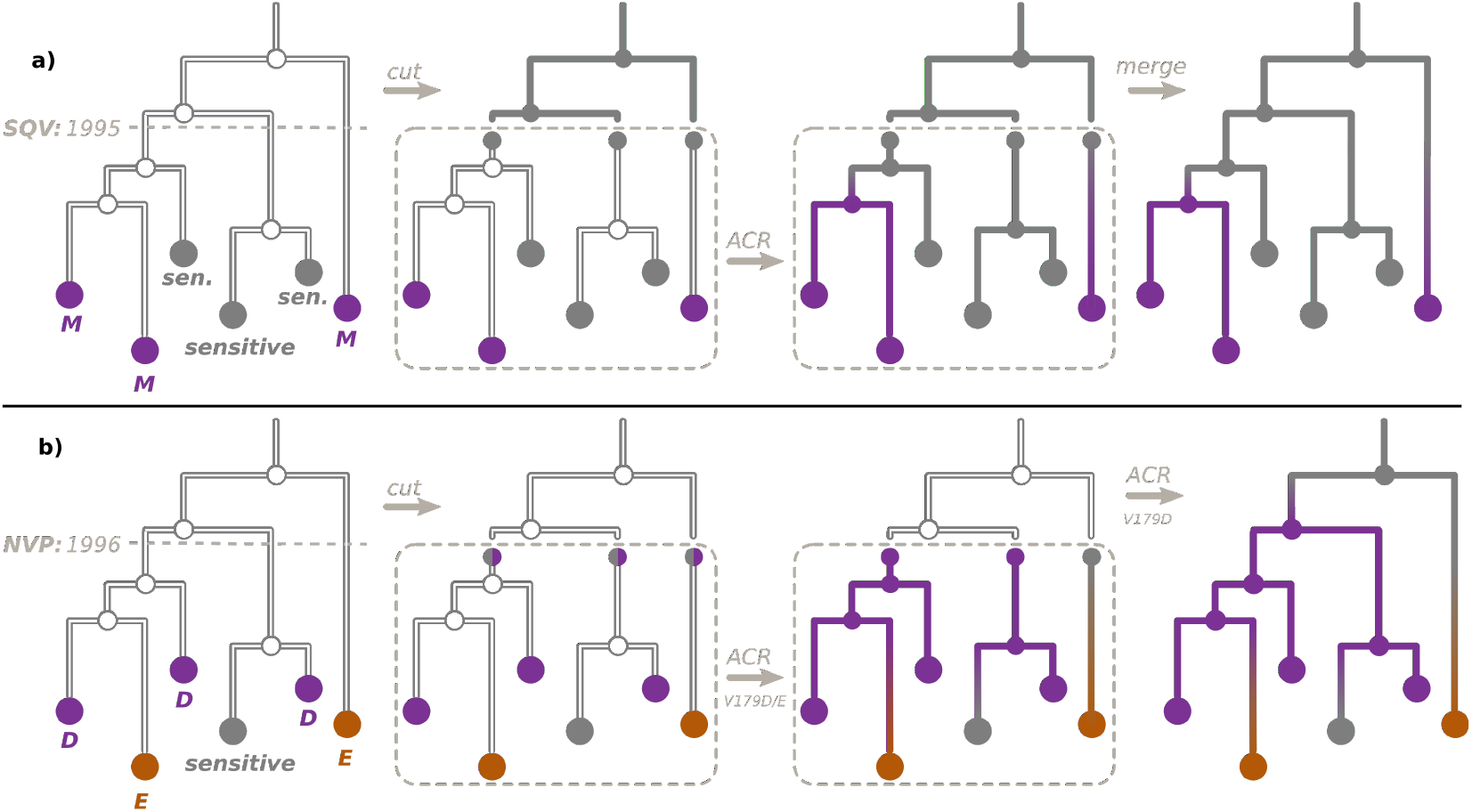
ACR for DRMs. a) To reconstruct the ancestral character states, resistant (violet, e.g. M) or sensitive (gray), for a non-polymorphic DRM (e.g. PR:L90M), we cut the time-scaled tree at the date of acceptance of the first ARV that can provoke this DRM (for PR:L90M, SQV accepted in 1995), as shown in the left panel. We hence obtain the pre-treatment-introduction tree (upper part of the tree) and a forest of post-treatment-introduction subtrees (bottom part). For the trees in the forest we then mark their roots as sensitive (middle left panel). We perform the ACR with PastML on the forest (middle right panel) and combine the results with the the all-sensitive annotation for the pre-treatment-introduction tree nodes (right panel). b) To reconstruct the ancestral character states for the DRM position RT:V179, corresponding to a polymorphic DRM RT:V179D (violet), but also to a non-polymorphic DRM RT:V179E (orange), we cut the time-scaled tree at the date of acceptance of the first ARV that can provoke RT:V179E (NVP, accepted in 1996), as shown in the left panel. For the trees in the after-1996 forest we then mark their roots as either sensitive (gray) or D (violet, middle left panel) and perform the ACR with PastML (middle right panel). We then extended this reconstruction to the before-1996 tree only for RT:V179D (right panel).

For two of the multiple-DRM positions (RT:T215 and RT:K219) all the corresponding DRMs were non-polymorphic and provoked by the same ARVs (the earliest accepted being zidovudine (AZT, accepted in 1987) for all of them). We therefore cut the tree as explained above, and reconstructed the ACR for D, F, S, Y or sensitive (for RT:T215), and for E, N, Q or sensitive (for RT:K219) on the after-1987 forest.

Finally, for RT:V179, the mutation RT:V179D was polymorphic, while RT:V179E was non-polymorphic (provoked by nevirapine, NVP, accepted in 1996). To reconstruct ancestral characters for RT:V179, we followed the procedure visualised in Figure 1b: First, we cut the tree at 1996, and reconstructed the ancestral characters (E, D or sensitive) on the after-1996 forest (the input states for the forest roots were sensitive or D). We then extended this reconstruction on the before-1996 tree only for RT:V179D (i.e., possible states: D or sensitive).

Once ACR was performed for all the DRM positions, we combined the predictions into a common table mapping node names to their states. A node state was sensitive if no DRM was reconstructed for this node at any position, otherwise the state was a combination of DRMs reconstructed for this node in separate DRM analyses (e.g. RT:K103N+RT:V106I if those DRMs were reconstructed as present for the node of interest while the others were reconstructed as absent). We visualized this combined result using the COPY method of PastML.

#### Transmitted versus acquired drug resistance

On a tree whose nodes are annotated with their DRM status, present (resistant) or absent (sensitive), we defined three configurations: transmitted drug resistance (TDR), acquired drug resistance (ADR), and DRM loss (see Figure 2).

**Figure 2.**
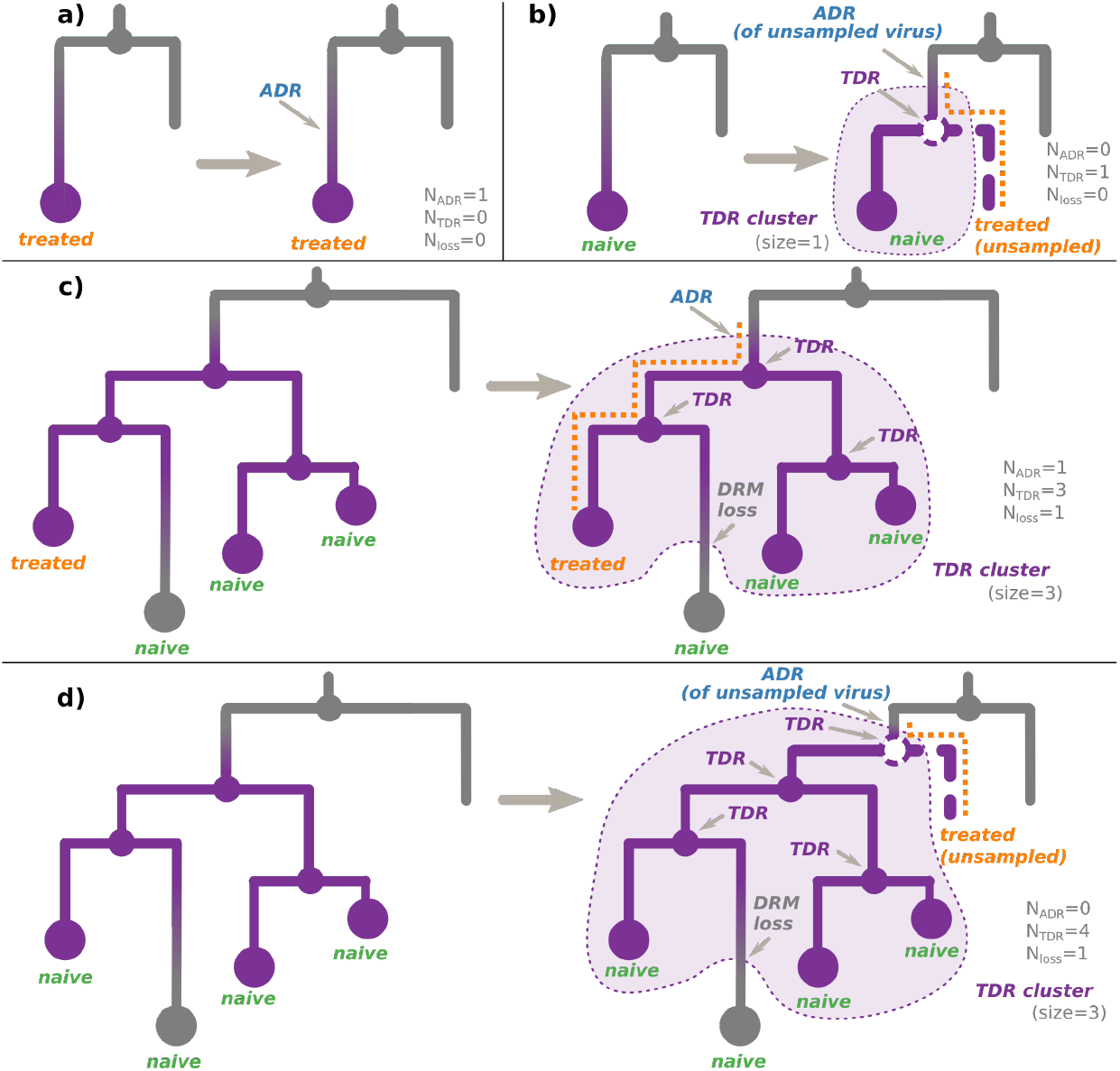
ADR and TDR scenarios. Each panel represents (left) a configuration observed in a tree whose nodes are annotated with their non-polymorphic DRM status (resistant nodes are violet, sensitive are gray), and (right) the most parsimonious transmission scenario (i.e. with the least number of events) leading to this configuration. The TDR clusters corresponding to the inferred scenarios are shown with violet background. **a)** The observed tree (left) contains a tip, corresponding to a sample of a resistant virus from a treatment-experienced individual, while its parent node (corresponding to a transmission) is sensitive. In the simplest scenario (right) the treatment-experienced individual’s virus acquired the DRM after the last observed transmission. **b)** The observed tree (left) contains a tip, corresponding to a sample of a resistant virus from a treatment-naive individual, while its parent node (corresponding to a transmission) is sensitive. The simplest scenario (right) includes a hidden transmission of a resistant virus from an unsampled treatment-experienced individual (dashed node and branch), whose virus previously acquired the DRM. **c)** The observed tree (left) contains one or several (here three) connected internal resistant nodes (corresponding to transmissions), leading to some treatment-naive tips (here three) and at least one treatment-experienced tip. Some of the tips might be sensitive (here one), while the others (here three) are resistant. In the simplest scenario (right) the treatment-experienced individual’s virus first acquired the DRM, then transmitted it to one (or several, here two) treatment-naive individuals, who might have further transmitted the resistant virus between them (here the transmission on the right). Some of the viruses might have eventually lost the DRM in the absence of drug-selective pressure (here the treatment-naive sensitive tip in the bottom). **d)** The observed tree (left) contains one or several (here three) connected internal resistant nodes (corresponding to transmissions), leading to only treatment-naive tips (here four). In the simplest scenario (right) an unsampled (and hence unobserved) treatment-experienced individual’s virus first acquired the DRM (before the oldest resistant internal node), then its host (dashed line) transmitted it (dashed node) to one (or several, here one) treatment-naive individuals of the observed cluster, who might have further transmitted the resistant virus between them (here all the three transmissions). Some of the viruses might have eventually lost the DRM in the absence of drug-selective pressure (here the treatment-naive sensitive tip in the bottom).

We defined ADR cases as parent-child node pairs, where the parent DRM status is sensitive, while the child DRM status is resistant.

We defined TDR cases inferred from the tree as either:

1. an internal node whose state was estimated as resistant (i.e. containing the DRM of interest, see Figure 2c,d). As the internal nodes of the tree roughly correspond to transmissions, such a node indicates a transmission of a resistant virus.
2. (for non-polymorphic mutations only) a hidden internal node between a node whose DRM status is resistant and its parent node whose DRM status is sensitive, if all the tips in the node’s subtree are treatment-naive. According to the treatment status and the fact that the mutation is non-polymorphic, the initial resistance could not be acquired through treatment pressure, and hence must have been transmitted from a patient who was not sampled (and does not appear in the tree, see Figure 2b,d).

Connected parts of the tree corresponding to TDR cases form TDR clusters (see Figure 2). We calculated their sizes as the numbers of resistant tips connected to each cluster. Note that if a TDR cluster subtree contains only treatment-naive patients, it implies that its root ADR event corresponds to an unsampled treated patient (see Figure 2b,d).

We define DRM loss cases as parent-child node pairs, where the parent DRM status is resistant, while the child DRM status is sensitive.

Using these configurations, we calculate the source of the DRM status of each tip in the tree as follows.

#### For non-polymorphic DRMs

1. For treatment-naive tips, the source of their DRM status is:
  - TDR if the tip is resistant (see Figure 2a,d);
  - TDR+DRM loss if the tip is sensitive and is involved in a DRM loss configuration (see Figure 2c,d);
  - transmission of a virus without the DRM if the above two cases do not apply.
2. For treatment-experienced tips, the source of their DRM status is:
  - ADR (+DRM loss if the tip is sensitive) for one of the treatment-experienced tips connected to a TDR cluster (see Figure 2c). The patient corresponding to this tip is assumed to be the source of the TDR cluster. The later DRM loss is possible if the treatment was changed to drugs that do not provoke the DRM in question. For other treated tips connected to this cluster, we assume that they received a resistant virus via TDR. Assuming their treatment was such that it could not provoke the DRM in question, they could later lose it (hence +DRM loss if they are sensitive);
  - ADR for a resistant tip not connected to a TDR cluster (Figure 2a);
  - transmission of a virus without the DRM if the above cases do not apply.
3. For the tips whose treatment status is unknown, we consider both cases (naive or resistant) with equal probabilities (0.5).

**For polymorphic DRMs** we do not consider the treatment status (as such DRMs could appear independently of treatment) and calculate the source of each tip’s DRM status as follows:

- ADR for a resistant tip not connected to a TDR cluster (as in Figure 2a, independently of the treatment status);
- ADR (+DRM loss if the tip is sensitive) for one of the tips connected to a TDR cluster (as in Figure 2c, independently of the treatment status). The individual corresponding to this tip is assumed to be the source of the TDR cluster. For other tips connected to this cluster, we assume that they received a resistant virus via TDR. They could later lose it (hence +DRM loss if they are sensitive);
- transmission of a virus without the DRM if the above cases do not apply.

We count the numbers of tip DRM status sources of each type (ADR: *N*_*ADR*_, TDR: *N*_*TDR*_, or loss: *N*_*loss*_ (see Appendix B for details) and report the results in Tables 2 and 3. We count all the identified DRM loss events, all the identified (observed and hidden) TDR events, and only those of the ADR events that are not at the root of naive-only TDR clusters, as the latter happened in unsampled treatment-experienced patients (see Figure 2b,d).

Note that *N*_*resistant tips*_ = *N*_*ADR*_ + *N*_*TDR*_ − *N*_*loss*_. For example, in Figure 2c, all the events correspond to observed tips, so we count one ADR, three TDR, and one DRM loss events: *N*_*resistant tips*_ = 3 = 1 + 3 − 1. Figure 2d represents a more complex case: We count one hidden TDR event (as it led to the resistance status of one of the observed tips) and three observed TDR events (leading to resistance statuses of other observed tips). We do not count the ADR event (as it corresponds to an unobserved patient, whose virus is not in our data set). We also count one DRM loss event, which led to one of the tips regaining its sensitive state. Hence *N*_*resistant tips*_ = 3; *N*_*ADR*_ = 0; *N*_*TDR*_ = 4; *N*_*loss*_ = 1; 3 = 0 + 4 − 1.

#### Times of DRM loss

We estimated the loss times for non-polymorphic DRMs, using survival analysis with an exponential (constant hazard) model (Weibull model with *β* = 1), implemented in Python3 package SurPyval (github.com/derrynknife/SurPyval, v0.10.10). This model takes as input observations about event durations and estimates the rate at which the event occurs. The input data might be left-, right- or interval-censored. Left-censored data represent times that are longer than the event occurrences, e.g. if the DRM loss occurred in exactly 2 years, but the observation was only made after 3 years, the 3-year duration represents a left-censored data point. Right-censored data represent times that are shorter than the event occurrences, e.g. if for the same DRM loss the only observation was made after 1 year (and observed no DRM loss yet), the 1-year duration represents a right-censored data point. Interval-censored data represents cases when both a left- and a right-censored data point are available, e.g. for the same DRM loss an interval-censored data might state that it occurred sometime between 1 and 3 years.

For each individual represented in our data set we extracted at most one data point for the loss survival analysis, as described below.

A right-censored data point represents the maximal observed duration during which a mutation loss did not occur. We extracted such points for the individuals who had several consecutive treatment-naive samples with the DRM of interest (and of the subtype of interest) in our metadata: we took the difference in sampling times of the last such sample and the first one.

A left-censored data point represents a duration that is longer than the mutation loss time. To estimate such a duration we needed to know not only (1) the time by which the individual’s virus lost the DRM, but also (2) the earliest time by which it could have acquired it (the difference making an upper limit on the loss duration). For (1) we used the time of the earliest sample without the DRM, provided it was preceded by samples with the DRM. For (2) we used either (2a) the time of the latest sample without the DRM preceding the aforementioned samples (where the DRM was present and then lost), if such sample existed in the metadata, or (2b) if the earliest metadata sample already had the DRM (which implies it corresponded to a resistant tip in the tree), the time of the tip’s most recent ancestral node whose status was sensitive (with marginal probability > 0.95).

For individuals for whom both a left- and a right-censored data point was present, we converted them to an interval-censored one.

We reported the resulting DRM loss time estimates (i.e. inverse of the loss rates) for non-polymorphic DRMs with at least 5 left-censored and 5 right-censored data points (interval-censored data points counted as both). We estimated confidence intervals (CIs) as the 2.5- and 97.5-percentiles of the loss times estimated on bootstrapped data points of the same size (with 1 000 repetitions).

## 3. Results

### 3.1. HIV in the UK

Antiretroviral therapy (ART) was introduced in the UK more than 30 years ago and transformed HIV from a fatal infection into a chronic, manageable condition [22–24]. It is accepted that successful ART results in an “undetectable” viral load which is protective from passing on the virus to others [25,26].

In the UK, a patient’s viral load is regularly monitored by the clinicians: Patients attend bi-annual or quarterly clinical visits, depending on how well they do on treatment. Moreover, the increase in viral load comes with symptoms (generally opportunistic infections that persist longer than they should). A suspicious increase from undetectable to detectable viral load (i.e. *viral rebound*) is the first sign of treatment failure.

In case of a viral rebound, the virus is sequenced to discriminate between *resistance* (presence of a known DRM) and *poor adherence* (failure without DRM, if a patient does not take the drugs regularly according to prescription). If resistance is the reason for a treatment failure, the treatment is changed.

Therefore, in the case of treatment failure, there is a window of opportunity for the virus to be transmitted: between the time the viral load increases to transmittable levels and the time when the clinician realizes it and changes treatment. The probability of transmission varies across patients, and depends on various factors [5].

The information collected from the HIV drug resistance tests carried out in the UK since 1996 is available in the UK HIV Drug Resistance Database. The database stores protease (PR) and reverse transcriptase (RT) sequences for about 50% of infected individuals in the UK.

### 3.2. UK HIV data set

We used the data from the UK HIV Drug Resistance Database containing samples from 1996 to 2016 to estimate transmission mechanisms for different common DRMs.

Out of 88 009 initial sequences obtained from the database, the majority were of subtypes B (58 569 sequences, 66.5%) and C (27 151 sequences, 30.1%), we also detected 8 D, 1 F, 2 G, and 3 K (< 0.0001%) sequences, and 2 276 potentially recombinant sequences (2.6%, in particular 494 A,B,G and 446 B,K-recombinants (0.5%)). We report these and other data set statistics in Table 1.

**Table 1.** Statistics on the B and C data sets. The “with DRM(s)” statistics count samples with at least one unambiguous resistant amino acid at any DRM position. Samples that contained either non-resistant or ambiguous amino acids at all DRM positions were considered as “without DRMs”. “p DRM(s)” stands for polymorphic DRMs, while “np DRMs” stands for non-polymorphic ones. Note that the same sequence might contain both p and np DRMs (at different positions).

|  |  | B | C |
| --- | --- | --- | --- |
| **total** |  | **58 569** | **27 151** |
| **filtered by patient** (first only, % of total) |  | **40 055** (68%) | **19 139** (70%) |
| – **without temporal outliers** (% of filtered) |  | **39 159** (99%) | **18 809** (98%) |
| – with DRM(s) (% of w/o outliers) |  | 12 300 (31%) | 5 148 (27%) |
| – w. 1 DRM (% of w/o outliers) |  | 7 257 (19%) | 3 174 (17%) |
| – w. $\geq 2$ DRMs (% of w/o outliers) | | 5 043 (13%) | 1 974 (10%) |
| – with np DRM(s) (% of w/o outliers) |  | 7 641 (20%) | 3 014 (16%) |
| – w. 1 np DRM (% of w/o outliers) |  | 3 852 (10%) | 1 496 (8%) |
| – w. $\geq 2$ np DRMs (% of w/o outliers) | | 3 789 (10%) | 1 518 (8%) |
| – with p DRM(s) (% of w/o outliers) |  | 5 740 (15%) | 2 673 (14%) |
| – w. 1 p DRM(s) (% of w/o outliers) |  | 5 416 (14%) | 2 538 (13%) |
| – w. $\geq 2$ p DRM(s) (% of w/o outliers) | | 324 (1%) | 135 (1%) |
| **Number of sequences** | **treatment-naïve** (% of w/o outliers) | **28 175** (72%) | **12 286** (65%) |
|  | – with DRM(s) (% of tr.-naïve) | 7 091 (25%) | 2 361 (19%) |
|  | – with np DRM(s) (% of tr.-naïve) | 3 364 (12%) | 829 (7%) |
|  | – with p DRM(s) (% of tr.-naïve) | 4 260 (15%) | 1 656 (13%) |
|  | **treatment-experienced** (% of w/o outliers) | **7 732** (20%) | **4 503** (24%) |
|  | – with DRM(s) (% of tr.-experienced) | 4 141 (54%) | 2 112 (47%) |
|  | – with np DRM(s) (% of tr.-experienced) | 3 618 (47%) | 1 730 (38%) |
|  | – with p DRM(s) (% of tr.-experienced) | 971 (13%) | 665 (15%) |
| **treatment-unknown** (% of w/o outliers) |  | **3 252** (8%) | **2 020** (11%) |
| **Root date** (95% CI) |  | **1965** ('59-'65) | **1944** ('29-'49) |
| **Mutation rate** (95% CI) $\cdot 10^{-3} \left[ \frac{\text{mutations}}{\text{site-year}} \right]$ | | **1.9** (1.8-1.9) | **1.4** (1.3-1.4) |
| **Phylogenetic diversity** = $\frac{\text{tree length}}{\text{number of branches}} \left[ \frac{\text{mutations}}{\text{site-branch}} \right]$ | | **0.014** | **0.019** |

**Table 2.**
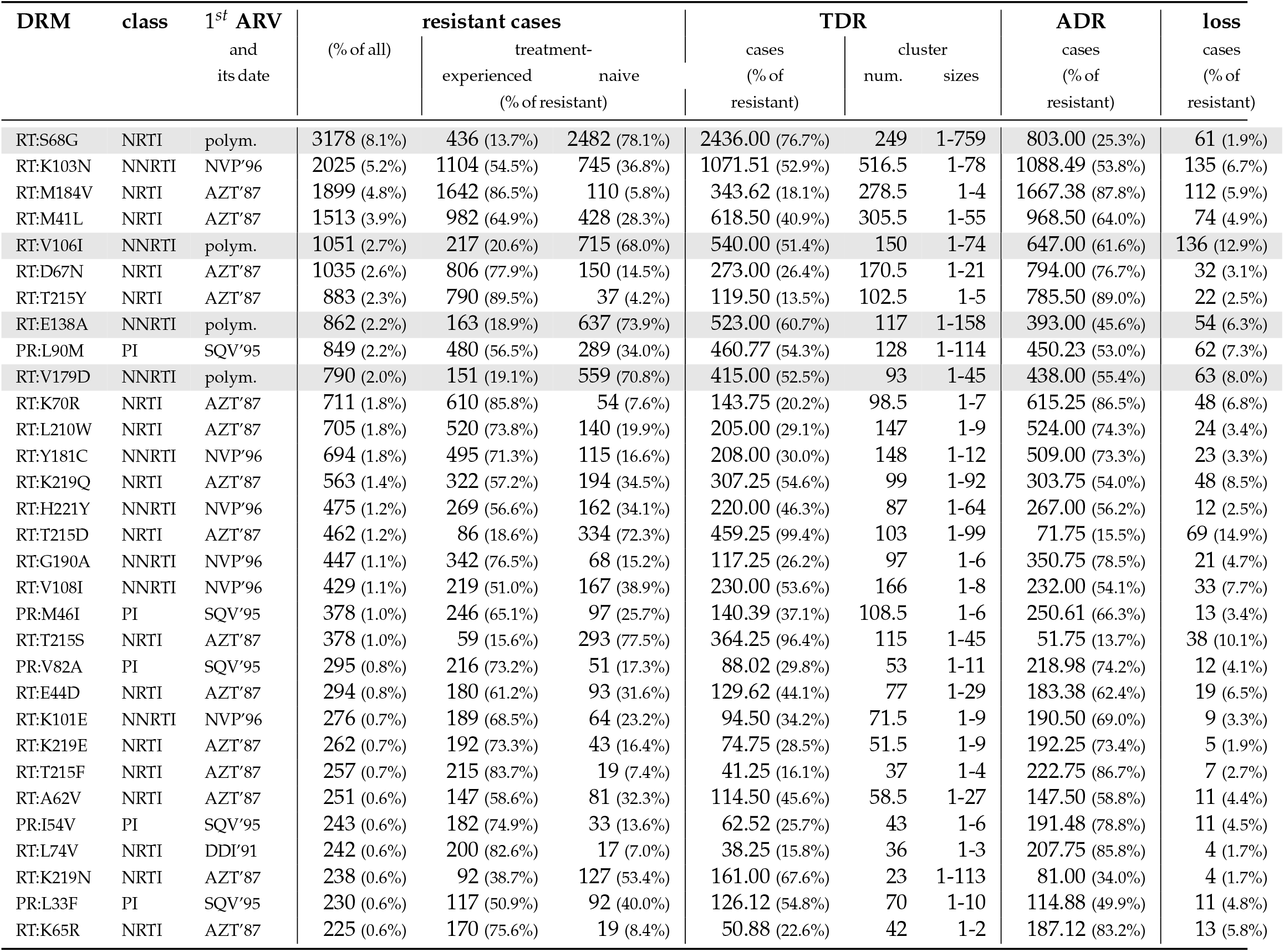
DRMs in B data set with prevalence > 0.5%. Polym. stands for polymorphic DRMs, for non-polymorphic DRMs the first ARV that could provoke it and its acceptance date are shown. *N*_*resistant cases*_ = *N*_*TDR*_ + *N*_*ADR*_ − *N*_*loss*_.

| DRM | class | 1<sup>st</sup> ARV<br>and<br>its date | (%) of all) | resistant cases |  | TDR |  |  | ADR<br>cases<br>(% of<br>resistant) | loss<br>cases<br>(% of<br>resistant) |
| --- | --- | --- | --- | --- | --- | --- | --- | --- | --- | --- |
|  |  |  |  | treatment-experienced | naive | cases<br>(% of<br>resistant) | cluster<br>num. | sizes |  |  |
|  |  |  |  | (% of resistant) |  |  |  |  |  |  |
| RT:S68G | NRTI | polym. | 3178 (8.1%) | 436 (13.7%) | 2482 (78.1%) | 2436.00 (76.7%) | 249 | 1-759 | 803.00 (25.3%) | 61 (1.9%) |
| RT:K103N | NNRTI | NVP'96 | 2025 (5.2%) | 1104 (54.5%) | 745 (36.8%) | 1071.51 (52.9%) | 516.5 | 1-78 | 1088.49 (53.8%) | 135 (6.7%) |
| RT:M184V | NRTI | AZT'87 | 1899 (4.8%) | 1642 (86.5%) | 110 (5.8%) | 343.62 (18.1%) | 278.5 | 1-4 | 1667.38 (87.8%) | 112 (5.9%) |
| RT:M41L | NRTI | AZT'87 | 1513 (3.9%) | 982 (64.9%) | 428 (28.3%) | 618.50 (40.9%) | 305.5 | 1-55 | 968.50 (64.0%) | 74 (4.9%) |
| RT:V106I | NNRTI | polym. | 1051 (2.7%) | 217 (20.6%) | 715 (68.0%) | 540.00 (51.4%) | 150 | 1-74 | 647.00 (61.6%) | 136 (12.9%) |
| RT:D67N | NRTI | AZT'87 | 1035 (2.6%) | 806 (77.9%) | 150 (14.5%) | 273.00 (26.4%) | 170.5 | 1-21 | 794.00 (76.7%) | 32 (3.1%) |
| RT:T215Y | NRTI | AZT'87 | 883 (2.3%) | 790 (89.5%) | 37 (4.2%) | 119.50 (13.5%) | 102.5 | 1-5 | 785.50 (89.0%) | 22 (2.5%) |
| RT:E138A | NNRTI | polym. | 862 (2.2%) | 163 (18.9%) | 637 (73.9%) | 523.00 (60.7%) | 117 | 1-158 | 393.00 (45.6%) | 54 (6.3%) |
| PR:L90M | PI | SQV'95 | 849 (2.2%) | 480 (56.5%) | 289 (34.0%) | 460.77 (54.3%) | 128 | 1-114 | 450.23 (53.0%) | 62 (7.3%) |
| RT:V179D | NNRTI | polym. | 790 (2.0%) | 151 (19.1%) | 559 (70.8%) | 415.00 (52.5%) | 93 | 1-45 | 438.00 (55.4%) | 63 (8.0%) |
| RT:K70R | NRTI | AZT'87 | 711 (1.8%) | 610 (85.8%) | 54 (7.6%) | 143.75 (20.2%) | 98.5 | 1-7 | 615.25 (86.5%) | 48 (6.8%) |
| RT:L210W | NRTI | AZT'87 | 705 (1.8%) | 520 (73.8%) | 140 (19.9%) | 205.00 (29.1%) | 147 | 1-9 | 524.00 (74.3%) | 24 (3.4%) |
| RT:Y181C | NNRTI | NVP'96 | 694 (1.8%) | 495 (71.3%) | 115 (16.6%) | 208.00 (30.0%) | 148 | 1-12 | 509.00 (73.3%) | 23 (3.3%) |
| RT:K219Q | NRTI | AZT'87 | 563 (1.4%) | 322 (57.2%) | 194 (34.5%) | 307.25 (54.6%) | 99 | 1-92 | 303.75 (54.0%) | 48 (8.5%) |
| RT:H221Y | NNRTI | NVP'96 | 475 (1.2%) | 269 (56.6%) | 162 (34.1%) | 220.00 (46.3%) | 87 | 1-64 | 267.00 (56.2%) | 12 (2.5%) |
| RT:T215D | NRTI | AZT'87 | 462 (1.2%) | 86 (18.6%) | 334 (72.3%) | 459.25 (99.4%) | 103 | 1-99 | 71.75 (15.5%) | 69 (14.9%) |
| RT:G190A | NNRTI | NVP'96 | 447 (1.1%) | 342 (76.5%) | 68 (15.2%) | 117.25 (26.2%) | 97 | 1-6 | 350.75 (78.5%) | 21 (4.7%) |
| RT:V108I | NNRTI | NVP'96 | 429 (1.1%) | 219 (51.0%) | 167 (38.9%) | 230.00 (53.6%) | 166 | 1-8 | 232.00 (54.1%) | 33 (7.7%) |
| PR:M46I | PI | SQV'95 | 378 (1.0%) | 246 (65.1%) | 97 (25.7%) | 140.39 (37.1%) | 108.5 | 1-6 | 250.61 (66.3%) | 13 (3.4%) |
| RT:T215S | NRTI | AZT'87 | 378 (1.0%) | 59 (15.6%) | 293 (77.5%) | 364.25 (96.4%) | 115 | 1-45 | 51.75 (13.7%) | 38 (10.1%) |
| PR:V82A | PI | SQV'95 | 295 (0.8%) | 216 (73.2%) | 51 (17.3%) | 88.02 (29.8%) | 53 | 1-11 | 218.98 (74.2%) | 12 (4.1%) |
| RT:E44D | NRTI | AZT'87 | 294 (0.8%) | 180 (61.2%) | 93 (31.6%) | 129.62 (44.1%) | 77 | 1-29 | 183.38 (62.4%) | 19 (6.5%) |
| RT:K101E | NNRTI | NVP'96 | 276 (0.7%) | 189 (68.5%) | 64 (23.2%) | 94.50 (34.2%) | 71.5 | 1-9 | 190.50 (69.0%) | 9 (3.3%) |
| RT:K219E | NRTI | AZT'87 | 262 (0.7%) | 192 (73.3%) | 43 (16.4%) | 74.75 (28.5%) | 51.5 | 1-9 | 192.25 (73.4%) | 5 (1.9%) |
| RT:T215F | NRTI | AZT'87 | 257 (0.7%) | 215 (83.7%) | 19 (7.4%) | 41.25 (16.1%) | 37 | 1-4 | 222.75 (86.7%) | 7 (2.7%) |
| RT:A62V | NRTI | AZT'87 | 251 (0.6%) | 147 (58.6%) | 81 (32.3%) | 114.50 (45.6%) | 58.5 | 1-27 | 147.50 (58.8%) | 11 (4.4%) |
| PR:I54V | PI | SQV'95 | 243 (0.6%) | 182 (74.9%) | 33 (13.6%) | 62.52 (25.7%) | 43 | 1-6 | 191.48 (78.8%) | 11 (4.5%) |
| RT:L74V | NRTI | DDI'91 | 242 (0.6%) | 200 (82.6%) | 17 (7.0%) | 38.25 (15.8%) | 36 | 1-3 | 207.75 (85.8%) | 4 (1.7%) |
| RT:K219N | NRTI | AZT'87 | 238 (0.6%) | 92 (38.7%) | 127 (53.4%) | 161.00 (67.6%) | 23 | 1-113 | 81.00 (34.0%) | 4 (1.7%) |
| PR:L33F | PI | SQV'95 | 230 (0.6%) | 117 (50.9%) | 92 (40.0%) | 126.12 (54.8%) | 70 | 1-10 | 114.88 (49.9%) | 11 (4.8%) |
| RT:K65R | NRTI | AZT'87 | 225 (0.6%) | 170 (75.6%) | 19 (8.4%) | 50.88 (22.6%) | 42 | 1-2 | 187.12 (83.2%) | 13 (5.8%) |

**Table 3.**
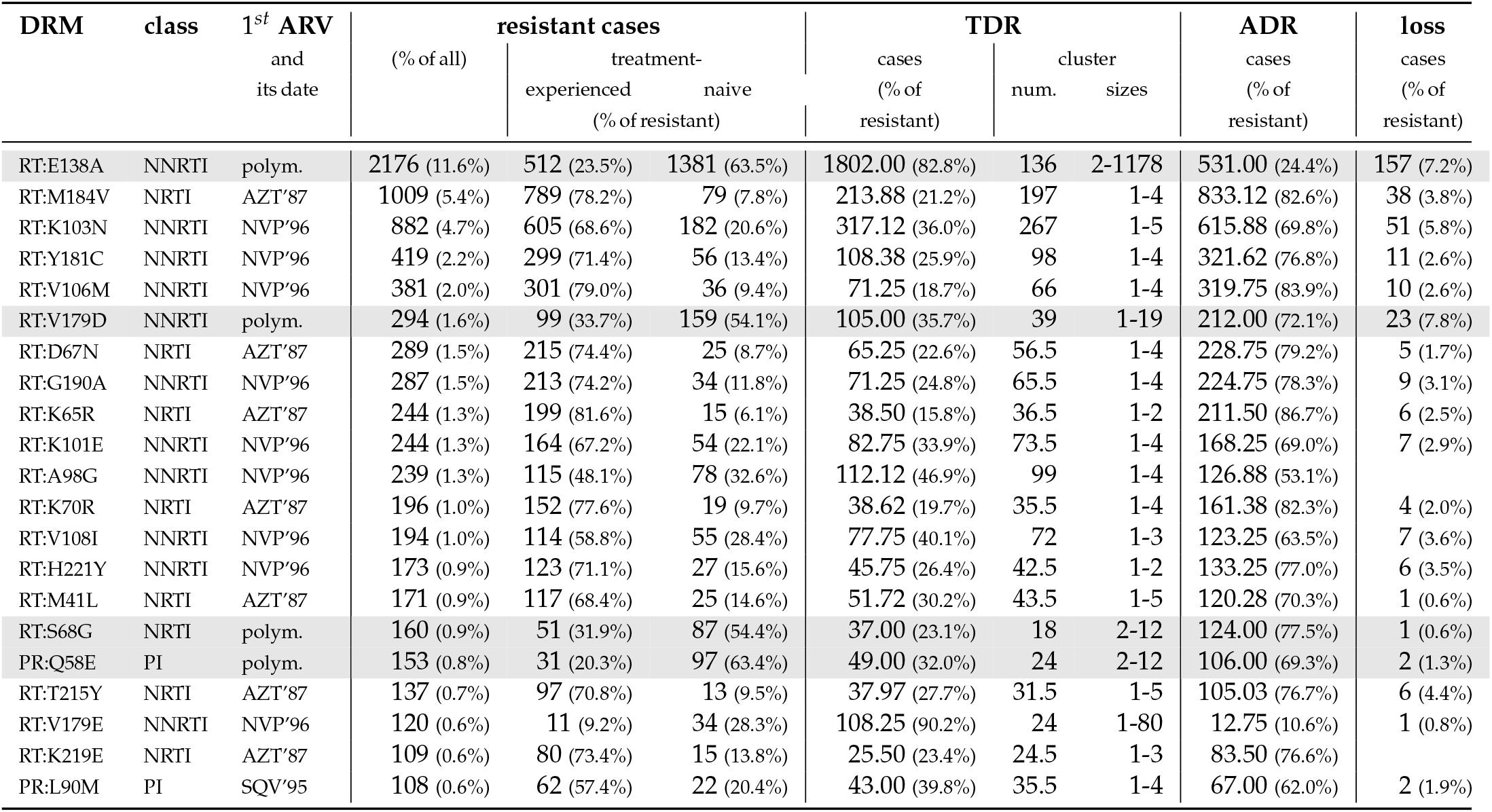
DRMs in C data set with prevalence > 0.5%. Polym. stands for polymorphic DRMs, for non-polymorphic DRMs the first ARV that could provoke it and its acceptance date are shown. *N*_*resistant cases*_ = *N*_*TDR*_ + *N*_*ADR*_ − *N*_*loss*_.

We focused our analysis on subtypes B and C, keeping the first sampled sequence for patients for whom multiple sequences were available. We hence obtained a 40 055-sequence data set for B, and a 19 139-sequence data set for C. We further filtered these data sets by removing temporal outliers (< 2% of sequences), as they could correspond to poorly sequenced samples or erroneous dates. The final data sets contained 39 159 sequences for B, and 18 809 for C.

We detected 161 DRMs found in at least one sequence of the B data set, and 146 DRMs for C. 31.4% of B and 27.4% of C sequences had at least one of these DRMs present, 18.5% of B and 16.9 % of C sequences had only one mutation, while the others had multiple DRMs present. While the subtypes B and C are different, as well as are the locations where these subtypes are most prevalent (African countries for C versus the UK and other European countries for B), we did not detect major differences in DRM distribution in the B and C data sets. Hence, while more C than B sequences correspond to imported cases, the UK health policies must play an important role on their DRM patterns, independently of the subtype. In a recent study Blassel *et al*. [27] compared DRMs in a UK and an African data sets. They reported that the median number of DRMs in resistant sequences differed between the two datasets (3 in the African sequences versus 1 in the UK sequences). In our case, there was no difference between B and C data sets if all DRMs were considered (median number of 1 DRM for both B and C data sets in resistant sequences); if we considered only non-polymorphic DRMs, a slight difference appeared (1 for B vs 2 for C). Detailed statistics on DRM number distributions are shown in Table A1. There was however a significant difference in the TDR distribution: more TDR could be suspected among the B samples (12% of treatment-naive sequences had non-polymorphic DRMs present, while in the C samples there were only 7% of such sequences).

### 3.3. Drug resistance analyses

We reconstructed time-scaled phylogenetic trees for B and C data sets and performed ancestral character reconstruction for each of the selected DRMs and positions to look at their transmission patterns. Consistently with what was previously reported in HIV-1 group M studies (of the *pol* gene [28] and of the full-genome [29]), we estimated a faster mutation rate (1.9 · 10^−3^ [mutations per site per year]) and a more recent root date (1965) for subtype B than for subtype C (1.4 · 10^−3^; 1944). More details on B and C datasets can be found in Table 1.

On the time-scaled trees we analyzed the transmission patterns of the DRMs found in at least 0.5% of sequences: 31 DRMs (on 26 different positions) for B and 21 (on 20 different positions) for C. The major drug resistance patterns found in B and C data sets are visualized in Figures 3 and 4. The statistics on these DRMs and their loss times are shown in Tables 2-4.

**Figure 3.**
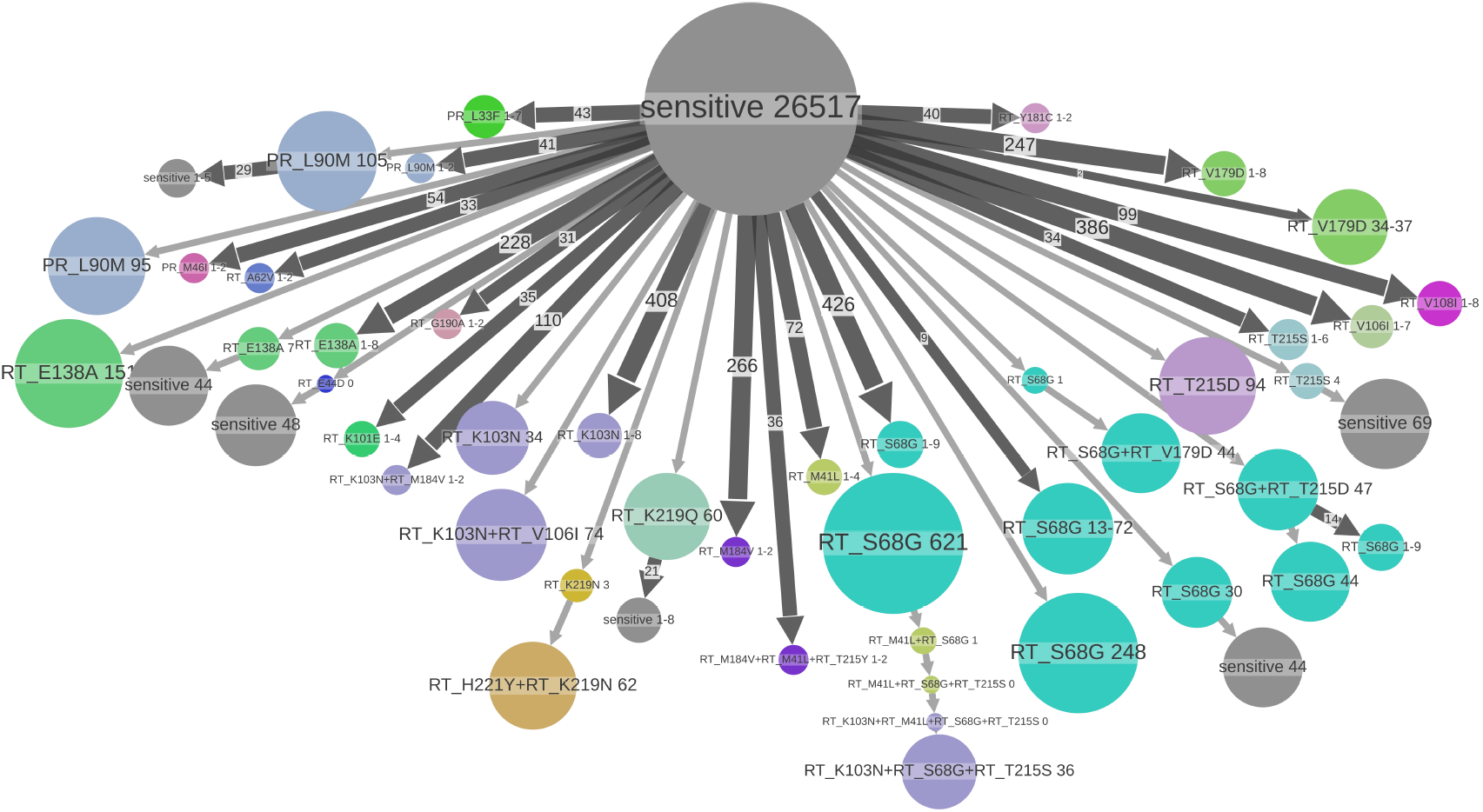
Major resistance patterns found in B data set. States of tree nodes are shown as labels, e.g.”PR_L90M” corresponds to the presence of DRM PR:L90M and absence of the other DRMs. The nodes are coloured by DRM found in them (if several DRMs are present, the colour of the (lexicographically) first DRM is used). The nodes with no DRM are coloured gray and labelled “sensitive”. The parts of the tree where no state change happens are clustered together into metanodes, their size corresponds to the number of samples (tips) they contain (shown in labels), e.g. “RT_K103N+RT_S68G+RT_T215S 36” (violet, on the bottom) corresponds to a transmitted resistance cluster containing 36 samples in the B data set, having three mutations. Configurations present several times are shown once and the number of occurrences is shown on the corresponding branch, e.g. a branch of size 247 leading to the metanode “RT_V179D 1-8” (salad green, on the top right) represents 247 cases of acquiring the mutation RT:V179D leading to small transmission clusters of sizes between 1 and 8. Configurations representing less than 34 samples are not shown to increase readability.

**Figure 4.**
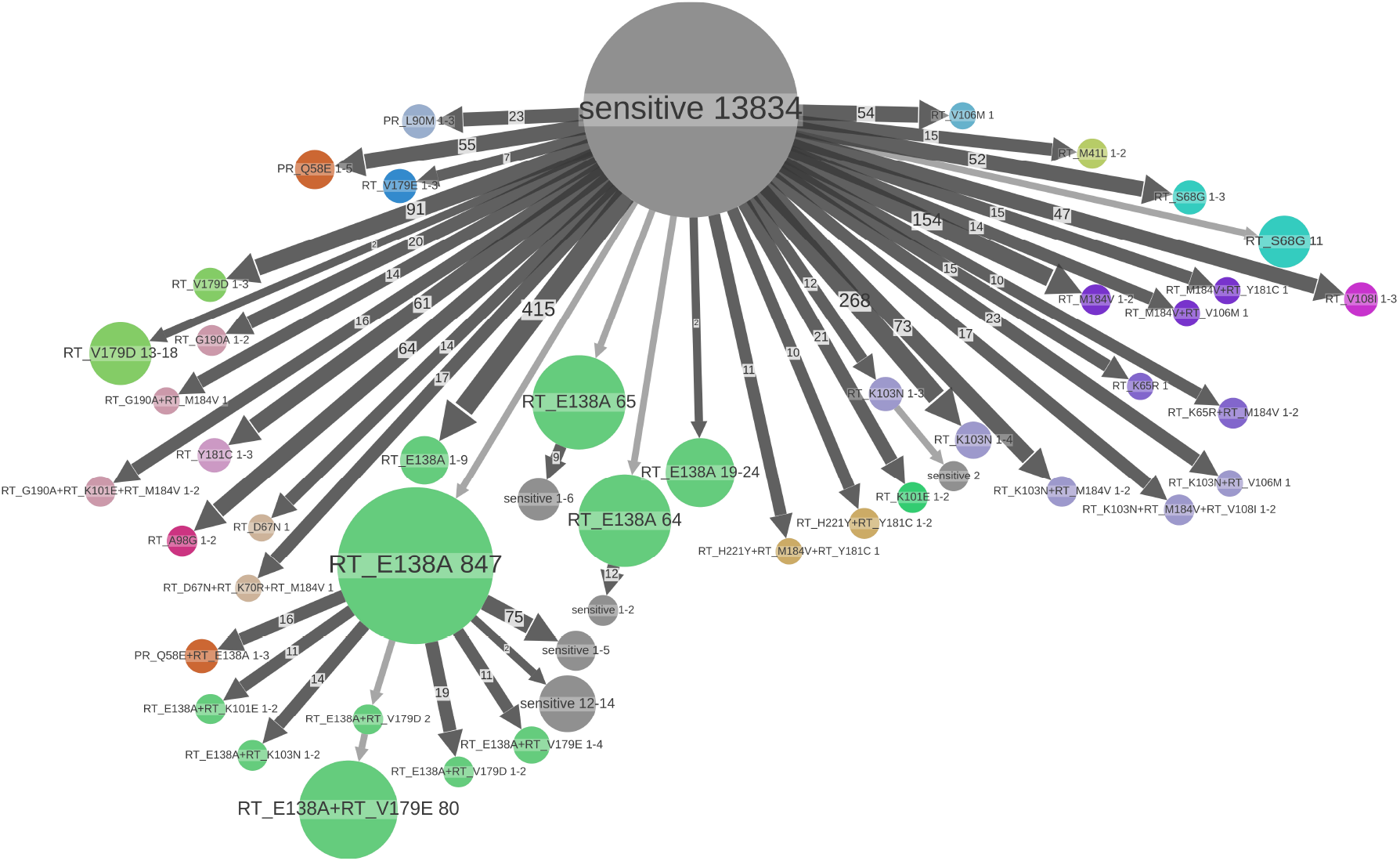
Major resistance patterns found in C data set. States of tree nodes are shown as labels, e.g. “RT_E138A” (salad green nodes in the middle) correspond to the presence of DRM RT:E138A and absence of the other DRMs. The nodes are coloured by DRM found in them (if several DRMs are present, the colour of the (lexicographically) first DRM is used). The nodes with no DRM found are coloured gray and labelled “sensitive”. The parts of the tree where no state change happens are clustered together into metanodes, their size corresponds to the number of samples (tips) they contain (shown in labels). Configurations present several times are shown once and the number of occurrences is shown on the corresponding branch, e.g. a branch of size 23 leading to the blue metanode “PR_L90M 1-3” (top left) represents 23 cases of acquiring the mutation PR:L90M leading to small resistance clusters of sizes 1–3. Configurations representing less than 11 samples are not shown to increase readability.

While some of the DRMs (e.g. RT:M184V) are comparably prevalent in B and C (4.8% of resistant cases vs 5.4%), others are very subtype-specific. For instance, the non-polymorphic mutation PR:L90M is present in 2.2% of B resistant cases and only in 0.6% of C. Another example is the mutations in position RT:106. In the B dataset the polymorphic DRM RT:V106I is present in 3.9% of resistant cases and is 30 times more prevalent than the non-polymorphic DRM RT:V106M, which was not selected for our analysis due to its low prevalence; while for the C data set we have the opposite distribution: RT:V106M is present in 2% of resistant cases and is 16 times more prevalent than RT:V106I, which was non selected for our analyses. More examples are given in Tables 2 and 3.

Using the metadata only, we can already see that there is a clear difference between polymorphic and non-polymorphic mutations. While the presence of most of the latter ones correlated with the treatment status (e.g. 86.5% of B sequences with the non-polymorphic mutation RT:M184V are from treatment-experienced patients), it is the opposite for the former, which are more prevalent in treatment-naive sequences (e.g. 78.1% of B sequences with RT:S68G are treatment-naive, see Table 2). Indeed, while the polymorphic DRMs can appear spontaneously, the non-polymorphic ones are selected by treatment, and carrying them often implies a fitness cost [9]. However, a few non-polymorphic DRM do not follow this pattern and are more prevalent in treatment-naive individuals: RT:T215D, RT:T215S, RT:K219N in B, and RT:V179E in C. RT:T215D/S area form of reversion and are are often developed in patients primarily infected with strains with RT:T215Y/F, and hence have a higher fitness [30]. It is further confirmed by our estimation of the loss times: the loss times of RT:T215D/S are long (9.3 and 6.8 years versus 1.1 and 1.8 years for RT:T215Y/F, see Table 4), which may explain their prevalence in treatment-naive patients. Similarly, we estimated a rather long loss time for RT:K219N (3.7 years). We did not have enough data to estimate the loss time of RT:V179E. While this mutation is generally considered as non-polymorphic [31], its natural presence in treatment-naive patients has been reported for the HIV-1 common recombinant form CRF55_01B [32].

**Table 4.**
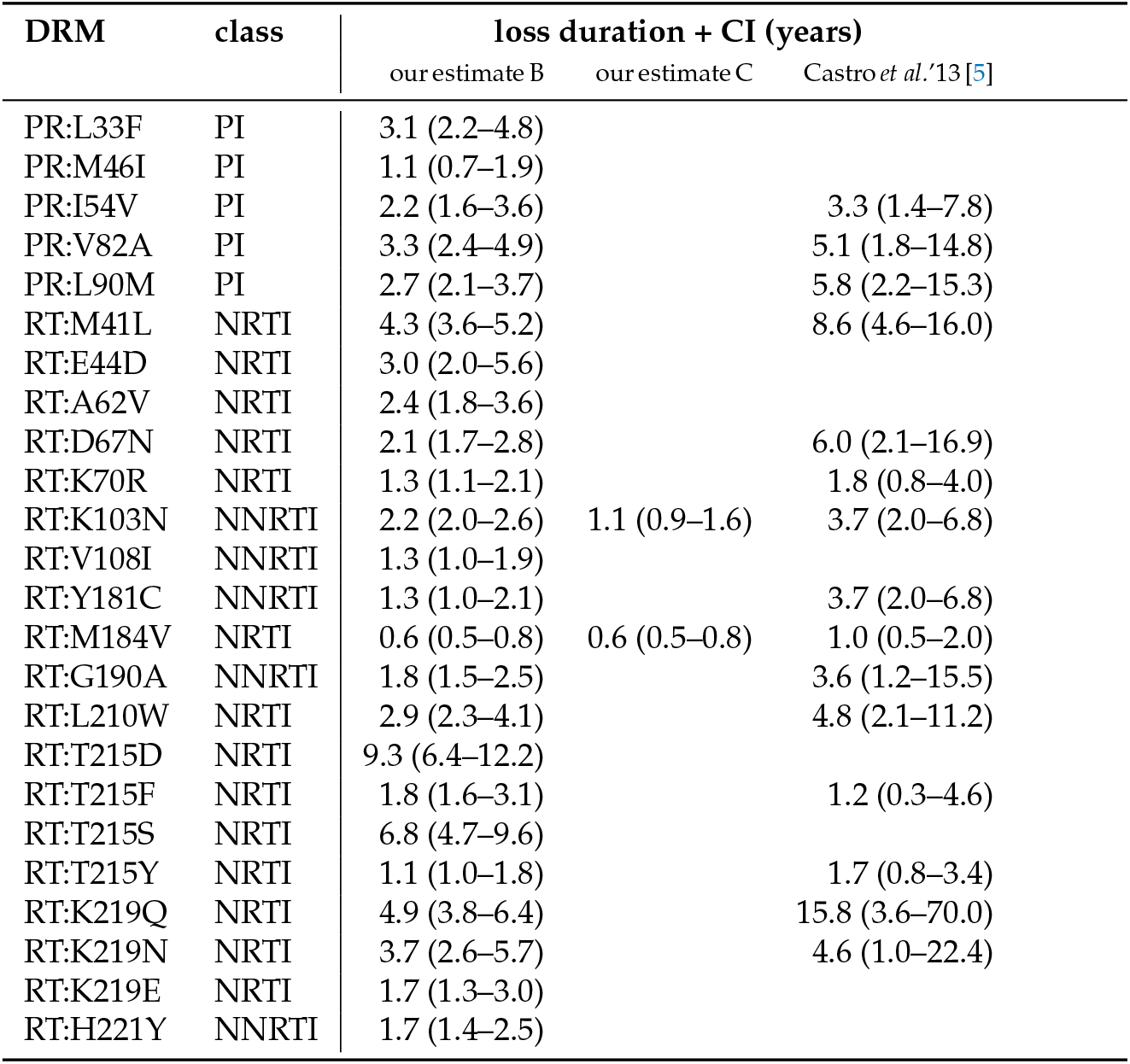
Loss times (with 95% CIs) for non-polymorphic DRMs found in B and C data sets with prevalence > 0.5% and at least 5 left- and 5 right-censored data points (the exact numbers of data points are shown in Table A3).

| DRM | class | loss duration + CI (years) |  |  |
| --- | --- | --- | --- | --- |
|  |  | our estimate B | our estimate C | Castro *et al.* '13 [5] |
| PR:L33F | PI | 3.1 (2.2–4.8) |  |  |
| PR:M46I | PI | 1.1 (0.7–1.9) |  |  |
| PR:I54V | PI | 2.2 (1.6–3.6) |  | 3.3 (1.4–7.8) |
| PR:V82A | PI | 3.3 (2.4–4.9) |  | 5.1 (1.8–14.8) |
| PR:L90M | PI | 2.7 (2.1–3.7) |  | 5.8 (2.2–15.3) |
| RT:M41L | NRTI | 4.3 (3.6–5.2) |  | 8.6 (4.6–16.0) |
| RT:E44D | NRTI | 3.0 (2.0–5.6) |  |  |
| RT:A62V | NRTI | 2.4 (1.8–3.6) |  |  |
| RT:D67N | NRTI | 2.1 (1.7–2.8) |  | 6.0 (2.1–16.9) |
| RT:K70R | NRTI | 1.3 (1.1–2.1) |  | 1.8 (0.8–4.0) |
| RT:K103N | NNRTI | 2.2 (2.0–2.6) | 1.1 (0.9–1.6) | 3.7 (2.0–6.8) |
| RT:V108I | NNRTI | 1.3 (1.0–1.9) |  |  |
| RT:Y181C | NNRTI | 1.3 (1.0–2.1) |  | 3.7 (2.0–6.8) |
| RT:M184V | NRTI | 0.6 (0.5–0.8) | 0.6 (0.5–0.8) | 1.0 (0.5–2.0) |
| RT:G190A | NNRTI | 1.8 (1.5–2.5) |  | 3.6 (1.2–15.5) |
| RT:L210W | NRTI | 2.9 (2.3–4.1) |  | 4.8 (2.1–11.2) |
| RT:T215D | NRTI | 9.3 (6.4–12.2) |  |  |
| RT:T215F | NRTI | 1.8 (1.6–3.1) |  | 1.2 (0.3–4.6) |
| RT:T215S | NRTI | 6.8 (4.7–9.6) |  |  |
| RT:T215Y | NRTI | 1.1 (1.0–1.8) |  | 1.7 (0.8–3.4) |
| RT:K219Q | NRTI | 4.9 (3.8–6.4) |  | 15.8 (3.6–70.0) |
| RT:K219N | NRTI | 3.7 (2.6–5.7) |  | 4.6 (1.0–22.4) |
| RT:K219E | NRTI | 1.7 (1.3–3.0) |  |  |
| RT:H221Y | NNRTI | 1.7 (1.4–2.5) |  |  |

Using the information from the tree, we refined the mutation statistics further, classifying resistant mutations sources into TDR vs ADR, and detecting DRM loss events (see Tables 2 and 3). C data set featured smaller TDR clusters (apart from the polymorphic mutation RT:E138A) than B. This could be explained by multiple introductions of subtype C into different regions of the UK and different risk groups, particularly from Africa via immigration, which is consistent with the higher diversity of the C strains observed in our data (C: 0.019 vs B: 0.014 [mutations per site per branch], Table 1) and with the dates of origin of the two UK sub-epidemics (C: 1944 vs B: 1965, Table 1).

A large size (e.g. 78 individuals in the B data set for RT:K103N) of some of the TDR clusters and a rather high proportion of TDR cases among the resistant ones (see Tables 2 and 3) is clinically problematic, as it means a high level of resistant strain transmission, leading to decrease of treatment choice on the population level.

We further analyzed each mutation position over time (see Supplementary Tables S1-S46) and found a common pattern: The proportion of resistant cases with respect to all cases decreases over time, however the proportion of resistant cases in treatment-naive individuals and, consistently, the proportion of TDR with respect to ADR, increases. This pattern is well illustrated by the mutations in position RT:215 (see Figure 5 and Table A2). However, there are exceptions with respect to the decrease in the proportion of resistant cases over time, especially among the polymorphic DRMs, consistent with the fact that they have little or no fitness cost associated with them. For the polymorphic mutation RT:E138A this proportions has been increasing from 2001 to mid-March 2016 (the last sampling time in our data): from 1.9% to 2.2% in the B data set, and from 8.3% to 11.6% in the C data set (Tables S8, S31). Similarly the proportion of resistant cases with polymorphic RT:S68G has been increasing from 4.6% in 2001 to 8.1% in 2016 in B, and from 0.3% to 0.9% in C (Tables S21, S41). The proportion of resistant cases with polymorphic RT:V106I has been increasing in B: from 2% in 2001 to 2.7% in 2016, while the proportion of non-polymorphic RT:V106M (similar to RT:V106I) in C seems to have stabilized at 2% over the last five sampling years (2011-2016, Tables S23, S43). The proportion of resistant cases with polymorphic RT:V179D has been increasing in B: from 1.3% in 2001 to 2% in 2016, so did the proportion of non-polymorphic RT:V179E (similar to RT:V179D) in C: from 0.1% in 2006 to 0.6% in 2016, while the proportion of RT:V179D has stayed stable (∼ 1.5%) over the last 10 sampling years (2006-2016, Tables S25, S45). Finally, the proportion of resistant cases with polymorphic PR:Q58E has been increasing in subtype C: from 0.6% in 2006 to 0.8% in 2016 (Table S28, we did not analyze it for B due to its low prevalence). These results clearly indicate that the spread of polymorphic DRMs should become a subject of particular surveillance.

**Figure 5.**
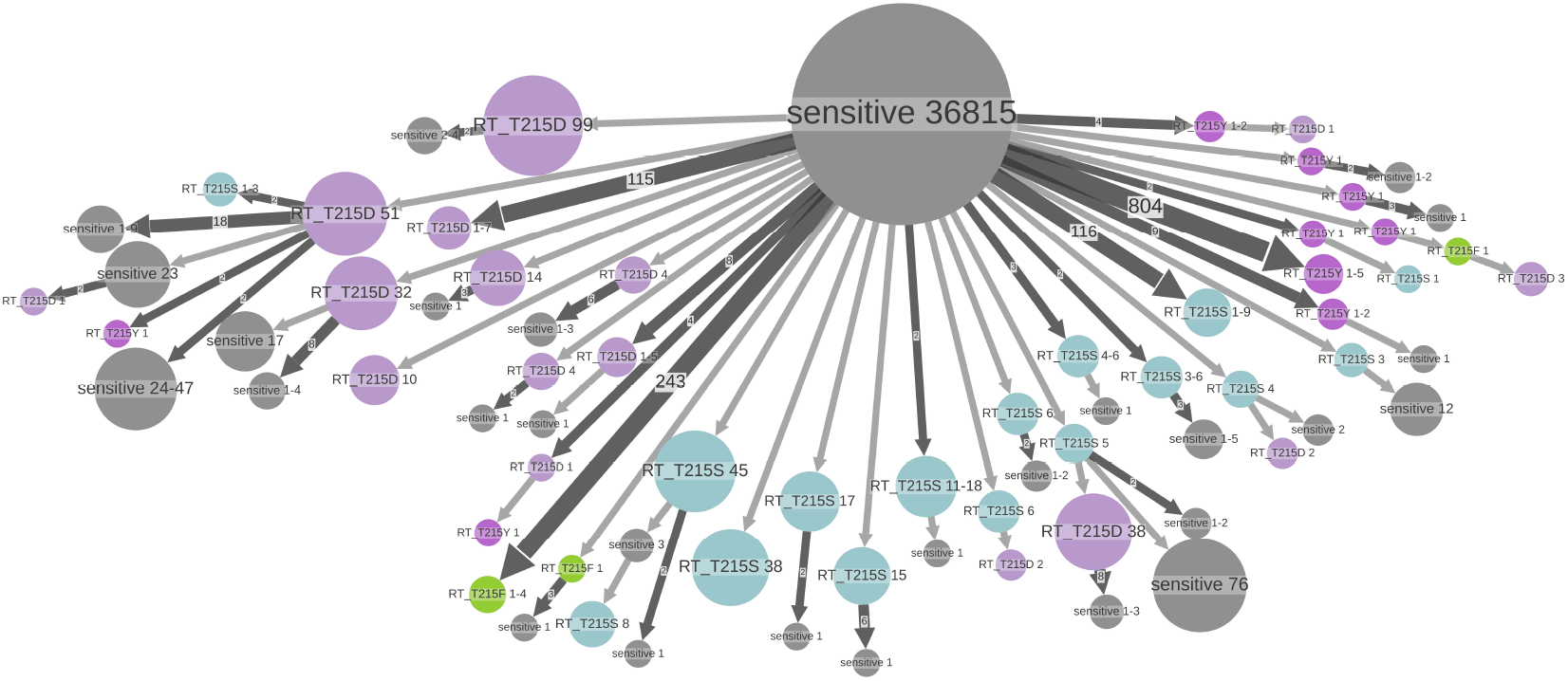
**DRMs with prevalence** > 0.5% **in position RT:215 in the B data set** (wildtype amino acid is T, non-polymorphic AZT-resistant mutations are D, F, S, and Y). ACR was performed for the RT position 215 with five possible states: D (lilac), F (salad green), S (light blue), Y (violet), and other (sensitive, gray). The parts of the tree where no state change happens are clustered together into metanodes, their size corresponds to the number of samples (tips) they contain (shown in labels), e.g. “RT_T215D 99” (lilac, top left) corresponds to a transmitted RT:T215D resistance cluster containing 99 samples in the B data set. Configurations present several times are shown once and the number of occurrences is shown on the corresponding branch, e.g. the branch of size 804 leading to the metanode “RT_T215Y 1-5” (violet, right) represents 804 cases of acquired RT:T215Y mutation leading to small transmission clusters of sizes between 1 and 5. Configurations representing less than 2 samples are not shown to increase readability.

#### 3.3.1. DRM loss times

We estimated the times of DRM loss for non-polymorphic DRMs in our data sets and compared them to the estimates previously reported by Castro *et al*. [5]. Castro *et al*. [5] analyzed 313 patients from the UK Drug Resistance database, who were treatment-naive and had a DRM present in their first resistance test (performed between 1997 and 2009), mixing all the subtypes and using survival analysis. We also used survival analysis, but had a larger data set, included the information not only from the metadata, but also from the tree, and analyzed the subtypes separately. Out results and the comparison are shown in Table 4. Overall, out estimates are compatible with those by Castro *et al*. [5]: the CIs of the two studies intersect for all the DRMs but RT:K103N in the C dataset. The difference for RT:K103N could be explained by the fact that Castro *et al*. [5] analyzed different subtypes together (though the majority of samples used were from B), while we performed a subtype-specific analysis: our estimate for RT:K103N on the B data set (2.0-2.6 years) is compatible with the one by Castro *et al*. [5] (2.0-6.8 years). Our CIs are systematically narrower than those of Castro *et al*. [5].

The visualization using ACR for the DRMs at the position RT:T215 (Figure 5) is consistent with the estimated loss patterns. For two of the mutations found in this position (RT:T215D and RT:T215S) the loss times are long (9.3 and 6.8 years respectively), which allows them to form large TDR clusters (up to 99 and 45 sampled respectively, left and bottom part of Figure 5). For the other two mutations found in this position (RT:T215F and RT:T215Y) the loss times (including potential reversions to D or S) are rather short (1.8 and 1.1 years), which prevents them from forming significant TDR clusters.

## 4. Discussion

We proposed fast maximum-likelihood ACR methods for investigation of drug resistance patterns in large sequence data sets. Their application to ∼ 40 000 subtype B and ∼ 20 000 subtype C sequences from the UK HIV Drug resistance database allowed us to investigate the trends in drug resistance patterns between 1996 and 2016 and to estimate the loss times for 25 common non-polymorphic DRMs.

An important advantage of our methods is their applicability to very large data sets (dozens of thousands of sequences). Previous studies had to face an uncomfortable choice between using more complex models on filtered data [9] or using less accurate (e.g. parsimony) approaches on full data sets [4]. Our approach uses a robust maximum likelihood framework, and permits the extraction of global drug resistant patterns from all the available data.

While the proportion of resistant cases in the UK seems to decrease with time, the proportion of resistant cases in treatment-naive individuals (hence acquired via TDR) is increasing. In addition, our results show that polymorphic DRMs obey to a different scheme, with an increase of both the proportion of resistant cases and TDR, and large resistance clusters. The TDR cases form resistance clusters, which are clearly identifiable on phylogenetic trees. Locating these clusters within the UK regions and cities, and among risk groups would be an important step in stopping drug resistance spread. The global trend that we observe in the UK is visible in other high-income countries (e.g. Switzerland [33], Italy [34] and Portugal [35]), but differs from, for example, West Africa, where the prevalence of multiple resistance in the population is a major concern [36]. Furthermore, detailed analyses in high-income countries indicate that a high level of ADR is more frequently observed in certain risk groups (e.g. African origin, unemployment, mental illness, among others, in Switzerland [37]) that require special surveillance to prevent treatment failure and HIV-1 transmission.

## Data Availability

All data produced are available online at https://github.com/evolbioinfo/HIV1-UK

## Author Contributions

Conceptualization, A.Z. and O.G.; methodology, A.Z. and O.G.; formal analysis, A.Z.; resources, D.D. and the UK HIV Drug Resistance Database & the Collaborative HIV, Anti-HIV Drug Resistance Network; writing–original draft preparation, A.Z. and O.G.; writing– review and editing, A.Z., O.G., and D.D.; visualization, A.Z.; supervision, O.G. All authors have read and agreed to the published version of the manuscript.

## Data Availability Statement

The HIV-1 sequences and the metadata (anonymized patient id and gender) used in this study, were obtained from the UK HIV Drug Resistance Database [14] in 2017. The visualizations of the ACR results produced in this study are available at github.com/evolbioinfo/HIV1-UK

## Acknowledgments

Authors thank Dr Stéphane Hué for valuable discussions on HIV spread in the UK.

## Abbreviations

The following abbreviations are used in this manuscript:

ADR: acquired drug resistance
ART: antiretroviral therapy
ARV: antiretroviral
AZT: zidovudine
CI: confidence interval
DDI: didanosine
DRM: drug resistance mutation
ETR: etravirine
MAP: maximum a posteriori
NFV: nelfinavir
NNRTI: non-nucleoside reverse transcriptase inhibitor
NRTI: nucleoside reverse transcriptase inhibitor
NVP: nevirapine
np DRM: non-polymorphic drug resistance mutation
PI: protease inhibitor
p DRM: polymorphic drug resistance mutation
PR: protease
RT: reverse transcriptase
SQV: saquinavir
TDF: tenofovir
TDR: transmitted drug resistance

## Appendix A Analysis pipelines

Snakemake [38] pipelines and ad hoc Python3 scripts used for the analyses described above are available on github.com/evolbioinfo/HIV1-UK. Along with the subtyping, tree reconstruction, dating and ACR tools mentioned above, we used goalign (v0.3.6) and gotree (v0.3.0b) [39] for basic sequence alignment and tree manipulations, as well as ETE3 framework [40] for basic tree manipulations (format conversion, pruning, etc.). Survival analysis was performed with Python3 package SurPyval (github.com/derrynknife/SurPyval). Sierra web service [20] for ARV and DRM detection was used via Python3 package sierrapy (github.com/hivdb/sierra-client).

## Appendix B

**Algorithm for counting** *N***_*ADR*_**, *N***_*TDR*_**, *N***_*loss*_ in a tree** 𝕋

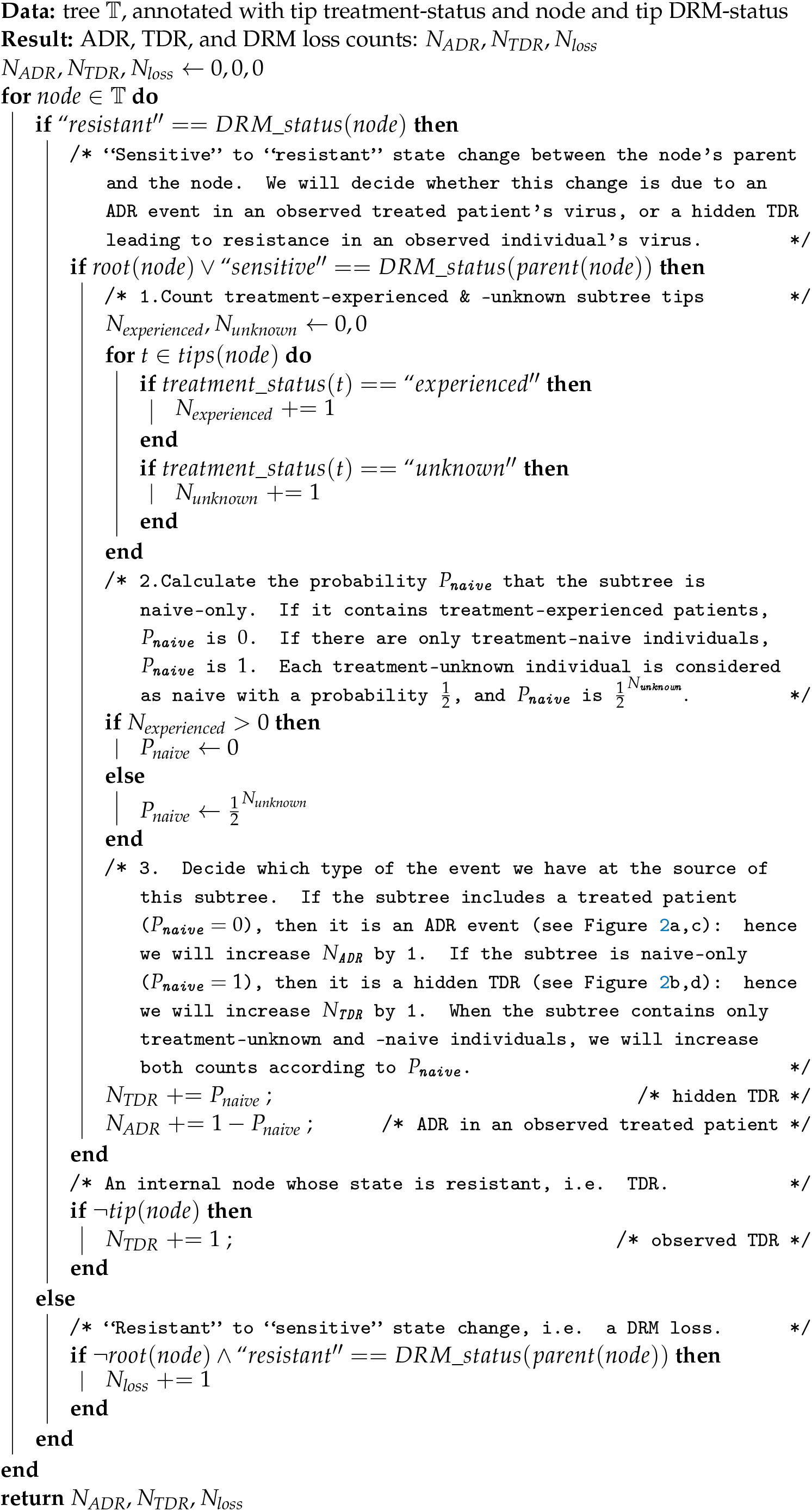

## Appendix C Additional Tables

**Table A1.**
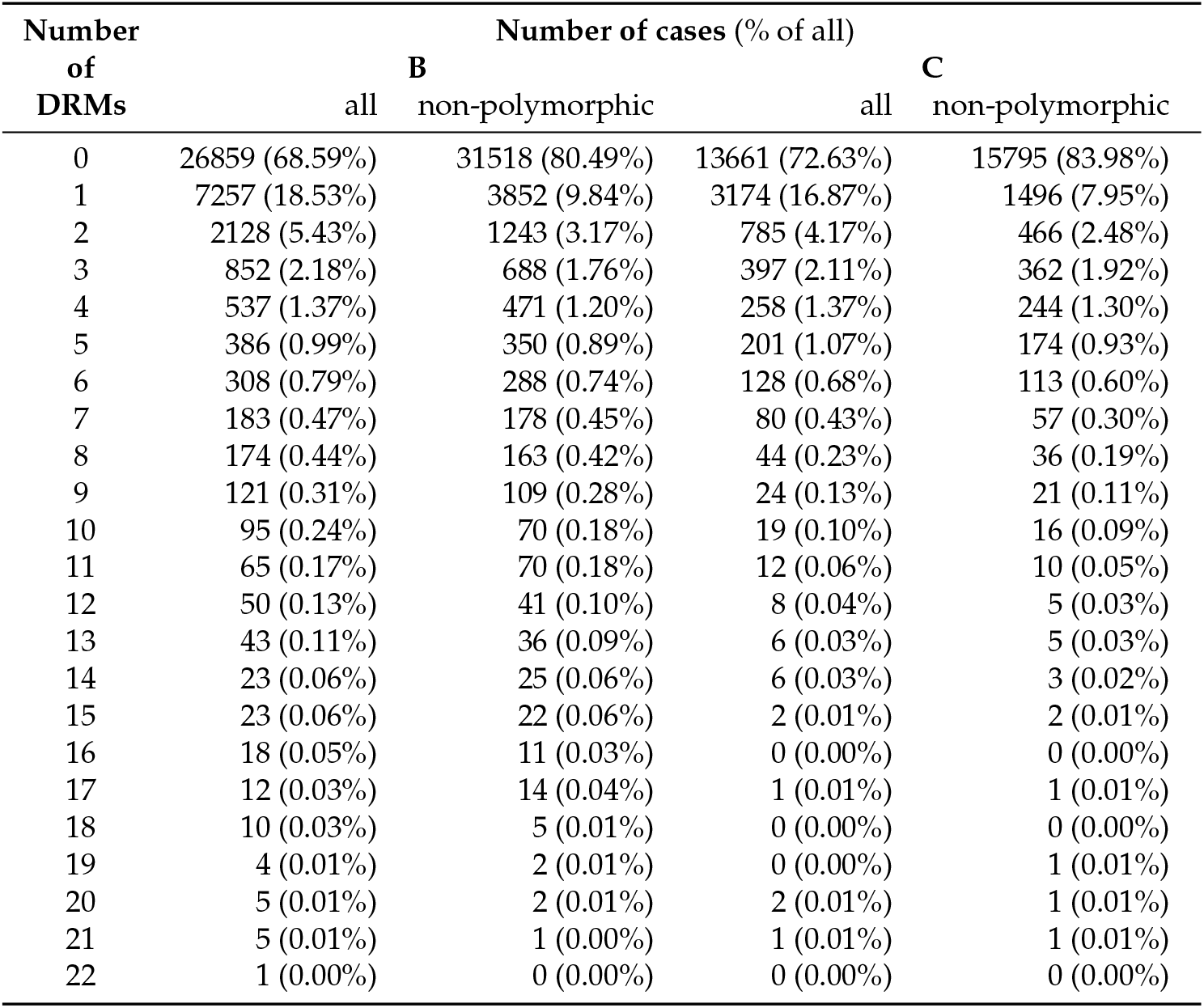
Resistant sequence counts in B and C data sets (after filtering by patient and temporal outlier removal).

| Number<br>of<br>DRMs | Number of cases (% of all) |  |  |  |
| --- | --- | --- | --- | --- |
|  | B |  | C |  |
|  | all | non-polymorphic | all | non-polymorphic |
| 0 | 26859 (68.59%) | 31518 (80.49%) | 13661 (72.63%) | 15795 (83.98%) |
| 1 | 7257 (18.53%) | 3852 (9.84%) | 3174 (16.87%) | 1496 (7.95%) |
| 2 | 2128 (5.43%) | 1243 (3.17%) | 785 (4.17%) | 466 (2.48%) |
| 3 | 852 (2.18%) | 688 (1.76%) | 397 (2.11%) | 362 (1.92%) |
| 4 | 537 (1.37%) | 471 (1.20%) | 258 (1.37%) | 244 (1.30%) |
| 5 | 386 (0.99%) | 350 (0.89%) | 201 (1.07%) | 174 (0.93%) |
| 6 | 308 (0.79%) | 288 (0.74%) | 128 (0.68%) | 113 (0.60%) |
| 7 | 183 (0.47%) | 178 (0.45%) | 80 (0.43%) | 57 (0.30%) |
| 8 | 174 (0.44%) | 163 (0.42%) | 44 (0.23%) | 36 (0.19%) |
| 9 | 121 (0.31%) | 109 (0.28%) | 24 (0.13%) | 21 (0.11%) |
| 10 | 95 (0.24%) | 70 (0.18%) | 19 (0.10%) | 16 (0.09%) |
| 11 | 65 (0.17%) | 70 (0.18%) | 12 (0.06%) | 10 (0.05%) |
| 12 | 50 (0.13%) | 41 (0.10%) | 8 (0.04%) | 5 (0.03%) |
| 13 | 43 (0.11%) | 36 (0.09%) | 6 (0.03%) | 5 (0.03%) |
| 14 | 23 (0.06%) | 25 (0.06%) | 6 (0.03%) | 3 (0.02%) |
| 15 | 23 (0.06%) | 22 (0.06%) | 2 (0.01%) | 2 (0.01%) |
| 16 | 18 (0.05%) | 11 (0.03%) | 0 (0.00%) | 0 (0.00%) |
| 17 | 12 (0.03%) | 14 (0.04%) | 1 (0.01%) | 1 (0.01%) |
| 18 | 10 (0.03%) | 5 (0.01%) | 0 (0.00%) | 0 (0.00%) |
| 19 | 4 (0.01%) | 2 (0.01%) | 0 (0.00%) | 1 (0.01%) |
| 20 | 5 (0.01%) | 2 (0.01%) | 2 (0.01%) | 1 (0.01%) |
| 21 | 5 (0.01%) | 1 (0.00%) | 1 (0.01%) | 1 (0.01%) |
| 22 | 1 (0.00%) | 0 (0.00%) | 0 (0.00%) | 0 (0.00%) |

**Table A2.** DRMs with prevalence > 0.5% found in position RT:T215 in B data set, and the evolution of their presence over time.

| date | total samples | DRM | resistant cases |  |  | TDR |  |  | ADR cases<br>(% of resistant) | loss cases<br>(% of resistant) |
| --- | --- | --- | --- | --- | --- | --- | --- | --- | --- | --- |
|  |  |  | (% of all) | treatment-experienced<br>(% of resistant) | naive<br>(% of resistant) | cases<br>(% of resistant) | cluster num. | sizes |  |  |
| 14-03-16 | 39159 | D | 462 (1.2%) | 86 (18.6%) | 334 (72.3%) | 459.25 (99.4%) | 103 | 1-99 | 71.75 (15.5%) | 69 (14.9%) |
|  |  | F | 257 (0.7%) | 215 (83.7%) | 19 (7.4%) | 41.25 (16.1%) | 37 | 1-4 | 222.75 (86.7%) | 7 (2.7%) |
|  |  | S | 378 (1.0%) | 59 (15.6%) | 293 (77.5%) | 364.25 (96.4%) | 115 | 1-45 | 51.75 (13.7%) | 38 (10.1%) |
|  |  | Y | 883 (2.3%) | 790 (89.5%) | 37 (4.2%) | 119.50 (13.5%) | 102.5 | 1-5 | 785.50 (89.0%) | 22 (2.5%) |
| 17-12-14 | 36258 | D | 441 (1.2%) | 83 (18.8%) | 322 (73.0%) | 437.25 (99.1%) | 102 | 1-88 | 70.75 (16.0%) | 67 (15.2%) |
|  |  | F | 256 (0.7%) | 214 (83.6%) | 19 (7.4%) | 41.25 (16.1%) | 37 | 1-4 | 221.75 (86.6%) | 7 (2.7%) |
|  |  | S | 358 (1.0%) | 53 (14.8%) | 284 (79.3%) | 348.75 (97.4%) | 109 | 1-44 | 47.25 (13.2%) | 38 (10.6%) |
|  |  | Y | 880 (2.4%) | 788 (89.5%) | 37 (4.2%) | 118.00 (13.4%) | 102 | 1-5 | 783.00 (89.0%) | 21 (2.4%) |
| 17-12-09 | 22540 | D | 314 (1.4%) | 61 (19.4%) | 234 (74.5%) | 309.50 (98.6%) | 83 | 1-47 | 60.50 (19.3%) | 56 (17.8%) |
|  |  | F | 241 (1.1%) | 204 (84.6%) | 17 (7.1%) | 36.50 (15.1%) | 33.5 | 1-4 | 209.50 (86.9%) | 5 (2.1%) |
|  |  | S | 226 (1.0%) | 34 (15.0%) | 178 (78.8%) | 222.00 (98.2%) | 75 | 1-21 | 35.00 (15.5%) | 31 (13.7%) |
|  |  | Y | 837 (3.7%) | 752 (89.8%) | 34 (4.1%) | 99.00 (11.8%) | 87 | 1-5 | 751.00 (89.7%) | 13 (1.6%) |
| 17-12-04 | 7511 | D | 123 (1.6%) | 31 (25.2%) | 85 (69.1%) | 109.50 (89.0%) | 45.5 | 1-21 | 37.50 (30.5%) | 24 (19.5%) |
|  |  | F | 185 (2.5%) | 160 (86.5%) | 14 (7.6%) | 24.00 (13.0%) | 24 | 1-2 | 163.00 (88.1%) | 2 (1.1%) |
|  |  | S | 70 (0.9%) | 17 (24.3%) | 43 (61.4%) | 63.75 (91.1%) | 34 | 1-8 | 22.25 (31.8%) | 16 (22.9%) |
|  |  | Y | 655 (8.7%) | 597 (91.1%) | 24 (3.7%) | 54.25 (8.3%) | 51 | 1-4 | 602.75 (92.0%) | 2 (0.3%) |
| 17-12-99 | 1576 | D | 28 (1.8%) | 9 (32.1%) | 17 (60.7%) | 20.00 (71.4%) | 14.5 | 1-2 | 10.00 (35.7%) | 2 (7.1%) |
|  |  | F | 42 (2.7%) | 41 (97.6%) | 1 (2.4%) | 2.00 (4.8%) | 2 | 1-2 | 40.00 (95.2%) |  |
|  |  | S | 9 (0.6%) | 3 (33.3%) | 4 (44.4%) | 5.00 (55.6%) | 4.5 | 1-2 | 4.00 (44.4%) |  |
|  |  | Y | 205 (13.0%) | 187 (91.2%) | 6 (2.9%) | 12.25 (6.0%) | 12 | 1-2 | 192.75 (94.0%) |  |
| 14-11-96 | 7 | D | 1 (14.3%) | 1 (100.0%) |  |  |  |  | 1.00 (100.0%) |  |
|  |  | F | 1 (14.3%) | 1 (100.0%) |  |  |  |  | 1.00 (100.0%) |  |
|  |  | S |  |  |  |  |  |  |  |  |
|  |  | Y | 2 (28.6%) | 2 (100.0%) |  |  |  |  | 2.00 (100.0%) |  |

**Table A3.** Numbers of data points available for loss time calculation for non-polymorphic DRMs found in B and C data sets (see Table 4).

| DRM | class | num. data points B |  |  | num. data points C |  |  |
| --- | --- | --- | --- | --- | --- | --- | --- |
|  |  | left- | right- | interval- | left- | right- | interval- |
|  |  |  | censored |  |  |  | censored |
| PR:L33F | PI | 36 | 17 | 0 |  |  |  |
| PR:M46I | PI | 95 | 8 | 1 |  |  |  |
| PR:I54V | PI | 77 | 7 | 4 |  |  |  |
| PR:V82A | PI | 80 | 17 | 1 |  |  |  |
| PR:L90M | PI | 153 | 35 | 0 |  |  |  |
| RT:M41L | NRTI | 238 | 68 | 4 |  |  |  |
| RT:E44D | NRTI | 50 | 9 | 1 |  |  |  |
| RT:A62V | NRTI | 68 | 14 | 0 |  |  |  |
| RT:D67N | NRTI | 231 | 25 | 4 |  |  |  |
| RT:K70R | NRTI | 216 | 4 | 2 |  |  |  |
| RT:K103N | NNRTI | 612 | 90 | 8 | 310 | 11 | 5 |
| RT:V108I | NNRTI | 148 | 9 | 2 |  |  |  |
| RT:Y181C | NNRTI | 264 | 9 | 5 |  |  |  |
| RT:M184V | NRTI | 825 | 7 | 3 | 408 | 4 | 7 |
| RT:G190A | NNRTI | 185 | 14 | 4 |  |  |  |
| RT:L210W | NRTI | 140 | 19 | 0 |  |  |  |
| RT:T215D | NRTI | 24 | 43 | 2 |  |  |  |
| RT:T215F | NRTI | 69 | 3 | 2 |  |  |  |
| RT:T215S | NRTI | 30 | 35 | 3 |  |  |  |
| RT:T215Y | NRTI | 223 | 5 | 1 |  |  |  |
| RT:K219E | NRTI | 72 | 8 | 1 |  |  |  |
| RT:K219Q | NRTI | 79 | 32 | 3 |  |  |  |
| RT:K219N | NRTI | 38 | 18 | 1 |  |  |  |
| RT:H221Y | NNRTI | 153 | 17 | 1 |  |  |  |

## Disclaimer/Publisher’s Note

The statements, opinions and data contained in all publications are solely those of the individual author(s) and contributor(s) and not of MDPI and/or the editor(s). MDPI and/or the editor(s) disclaim responsibility for any injury to people or property resulting from any ideas, methods, instructions or products referred to in the content.

