## Supplemental Tables for "Modelling drug resistance emergence and transmission in HIV-1 in the UK"

### 1. Resistance statistics over time by DRM

#### *1.1. Subtype B*

**Table S1.** DRMs with prevalence > 0.5% found in position PR:I54 in B data set, and the evolution of their presence over time.

| date | total samples | DRM | resistant cases (% of all) | resistant cases: treatment-experienced (% of resistant) | resistant cases: treatment-naive (% of resistant) | TDR cases (% of resistant) | TDR cluster num. | TDR cluster sizes | ADR cases (% of resistant) | loss cases (% of resistant) |
|---|---|---|---|---|---|---|---|---|---|---|
| 14-03-16 | 39159 | V | 243 (0.6%) | 182 (74.9%) | 33 (13.6%) | 62.52 (25.7%) | 43 | 1-6 | 191.48 (78.8%) | 11 (4.5%) |
| 17-12-11 | 28011 | V | 231 (0.8%) | 177 (76.6%) | 27 (11.7%) | 55.02 (23.8%) | 39 | 1-6 | 184.98 (80.1%) | 9 (3.9%) |
| 17-12-06 | 13280 | V | 190 (1.4%) | 152 (80.0%) | 20 (10.5%) | 36.12 (19.0%) | 29 | 1-3 | 158.88 (83.6%) | 5 (2.6%) |
| 17-12-01 | 3195 | V | 82 (2.6%) | 74 (90.2%) | 4 (4.9%) | 6.00 (7.3%) | 6 | 1-1 | 76.00 (92.7%) | |
| 14-11-96 | 7 | V | | | | | | | | |

**Table S2.** DRMs with prevalence > 0.5% found in position PR:L33 in B data set, and the evolution of their presence over time.

| date | total samples | DRM | resistant cases (% of all) | resistant cases: treatment-experienced (% of resistant) | resistant cases: treatment-naive (% of resistant) | TDR cases (% of resistant) | TDR cluster num. | TDR cluster sizes | ADR cases (% of resistant) | loss cases (% of resistant) |
|---|---|---|---|---|---|---|---|---|---|---|
| 14-03-16 | 39159 | F | 230 (0.6%) | 117 (50.9%) | 92 (40.0%) | 126.12 (54.8%) | 70 | 1-10 | 114.88 (49.9%) | 11 (4.8%) |
| 17-12-11 | 28011 | F | 191 (0.7%) | 104 (54.5%) | 71 (37.2%) | 95.75 (50.1%) | 53.5 | 1-9 | 102.25 (53.5%) | 7 (3.7%) |
| 17-12-06 | 13280 | F | 111 (0.8%) | 72 (64.9%) | 28 (25.2%) | 38.50 (34.7%) | 27.5 | 1-5 | 76.50 (68.9%) | 4 (3.6%) |
| 17-12-01 | 3195 | F | 26 (0.8%) | 22 (84.6%) | 2 (7.7%) | 3.00 (11.5%) | 3 | 1-1 | 23.00 (88.5%) | |
| 14-11-96 | 7 | F | | | | | | | | |

**Table S3.** DRMs with prevalence > 0.5% found in position PR:L90 in B data set, and the evolution of their presence over time.

| date | total samples | DRM | resistant cases (% of all) | resistant cases: treatment-experienced (% of resistant) | resistant cases: treatment-naive (% of resistant) | TDR cases (% of resistant) | TDR cluster num. | TDR cluster sizes | ADR cases (% of resistant) | loss cases (% of resistant) |
|---|---|---|---|---|---|---|---|---|---|---|
| 14-03-16 | 39159 | M | 849 (2.2%) | 480 (56.5%) | 289 (34.0%) | 460.77 (54.3%) | 128 | 1-114 | 450.23 (53.0%) | 62 (7.3%) |
| 17-12-11 | 28011 | M | 709 (2.5%) | 449 (63.3%) | 196 (27.6%) | 318.52 (44.9%) | 109 | 1-71 | 435.48 (61.4%) | 45 (6.3%) |
| 17-12-06 | 13280 | M | 513 (3.9%) | 390 (76.0%) | 88 (17.2%) | 137.62 (26.8%) | 81 | 1-25 | 386.38 (75.3%) | 11 (2.1%) |
| 17-12-01 | 3195 | M | 206 (6.4%) | 178 (86.4%) | 16 (7.8%) | 25.00 (12.1%) | 20 | 1-5 | 182.00 (88.3%) | 1 (0.5%) |
| 14-11-96 | 7 | M | | | | | | | | |

**Table S4.** DRMs with prevalence > 0.5% found in position PR:M46 in B data set, and the evolution of their presence over time.

| date | total samples | DRM | resistant cases (% of all) | resistant cases: treatment-experienced (% of resistant) | resistant cases: naive (% of resistant) | TDR cases (% of resistant) | TDR cluster num. | TDR cluster sizes | ADR cases (% of resistant) | loss cases (% of resistant) |
|---|---|---|---|---|---|---|---|---|---|---|
| 14-03-16 | 39159 | I | 378 (1.0%) | 246 (65.1%) | 97 (25.7%) | 140.39 (37.1%) | 108.5 | 1-6 | 250.61 (66.3%) | 13 (3.4%) |
| 17-12-11 | 28011 | I | 324 (1.2%) | 232 (71.6%) | 65 (20.1%) | 95.52 (29.5%) | 75.5 | 1-6 | 235.48 (72.7%) | 7 (2.2%) |
| 17-12-06 | 13280 | I | 238 (1.8%) | 191 (80.3%) | 30 (12.6%) | 48.12 (20.2%) | 41 | 1-4 | 194.88 (81.9%) | 5 (2.1%) |
| 17-12-01 | 3195 | I | 101 (3.2%) | 91 (90.1%) | 6 (5.9%) | 8.00 (7.9%) | 8 | 1-1 | 93.00 (92.1%) | |
| 14-11-96 | 7 | I | 1 (14.3%) | 1 (100.0%) | | | | | 1.00 (100.0%) | |

**Table S5.** DRMs with prevalence > 0.5% found in position PR:V82 in B data set, and the evolution of their presence over time.

| date | total samples | DRM | resistant cases (% of all) | resistant cases: treatment-experienced (% of resistant) | resistant cases: naive (% of resistant) | TDR cases (% of resistant) | TDR cluster num. | TDR cluster sizes | ADR cases (% of resistant) | loss cases (% of resistant) |
|---|---|---|---|---|---|---|---|---|---|---|
| 14-03-16 | 39159 | A | 295 (0.8%) | 216 (73.2%) | 51 (17.3%) | 88.02 (29.8%) | 53 | 1-11 | 218.98 (74.2%) | 12 (4.1%) |
| 17-12-11 | 28011 | A | 276 (1.0%) | 210 (76.1%) | 42 (15.2%) | 74.52 (27.0%) | 48 | 1-10 | 212.48 (77.0%) | 11 (4.0%) |
| 17-12-06 | 13280 | A | 220 (1.7%) | 179 (81.4%) | 25 (11.4%) | 42.12 (19.1%) | 33 | 1-3 | 183.88 (83.6%) | 6 (2.7%) |
| 17-12-01 | 3195 | A | 99 (3.1%) | 92 (92.9%) | 3 (3.0%) | 6.00 (6.1%) | 6 | 1-2 | 93.00 (93.9%) | |
| 14-11-96 | 7 | A | | | | | | | | |

**Table S6.** DRMs with prevalence > 0.5% found in position RT:A62 in B data set, and the evolution of their presence over time.

| date | total samples | DRM | resistant cases (% of all) | resistant cases: treatment-experienced (% of resistant) | resistant cases: naive (% of resistant) | TDR cases (% of resistant) | TDR cluster num. | TDR cluster sizes | ADR cases (% of resistant) | loss cases (% of resistant) |
|---|---|---|---|---|---|---|---|---|---|---|
| 14-03-16 | 39159 | V | 251 (0.6%) | 147 (58.6%) | 81 (32.3%) | 114.50 (45.6%) | 58.5 | 1-27 | 147.50 (58.8%) | 11 (4.4%) |
| 17-12-11 | 28011 | V | 208 (0.7%) | 134 (64.4%) | 57 (27.4%) | 80.00 (38.5%) | 42 | 1-16 | 134.00 (64.4%) | 6 (2.9%) |
| 17-12-06 | 13280 | V | 136 (1.0%) | 109 (80.1%) | 17 (12.5%) | 28.50 (21.0%) | 18.5 | 1-8 | 108.50 (79.8%) | 1 (0.7%) |
| 17-12-01 | 3195 | V | 55 (1.7%) | 45 (81.8%) | 6 (10.9%) | 11.00 (20.0%) | 8 | 1-4 | 44.00 (80.0%) | |
| 14-11-96 | 7 | V | | | | | | | | |

#### *1.2. Subtype C*

**Table S7.** DRMs with prevalence > 0.5% found in position RT:D67 in B data set, and the evolution of their presence over time.

| date | total samples | DRM | resistant cases (% of all) | resistant cases: treatment-experienced (% of resistant) | resistant cases: naive (% of resistant) | TDR cases (% of resistant) | TDR cluster num. | TDR cluster sizes | ADR cases (% of resistant) | loss cases (% of resistant) |
|---|---|---|---|---|---|---|---|---|---|---|
| 14-03-16 | 39159 | N | 1035 (2.6%) | 806 (77.9%) | 150 (14.5%) | 273.00 (26.4%) | 170.5 | 1-21 | 794.00 (76.7%) | 32 (3.1%) |
| 17-12-11 | 28011 | N | 965 (3.4%) | 772 (80.0%) | 126 (13.1%) | 222.25 (23.0%) | 144 | 1-20 | 766.75 (79.5%) | 24 (2.5%) |
| 17-12-06 | 13280 | N | 801 (6.0%) | 670 (83.6%) | 76 (9.5%) | 139.50 (17.4%) | 105.5 | 1-10 | 676.50 (84.5%) | 15 (1.9%) |
| 17-12-01 | 3195 | N | 364 (11.4%) | 329 (90.4%) | 20 (5.5%) | 32.50 (8.9%) | 31.5 | 1-2 | 332.50 (91.3%) | 1 (0.3%) |
| 14-11-96 | 7 | N | 4 (57.1%) | 4 (100.0%) | | | | | 4.00 (100.0%) | |

**Table S8.** DRMs with prevalence > 0.5% found in position RT:E138 in B data set, and the evolution of their presence over time.

| date | total samples | DRM | resistant cases (% of all) | resistant cases: treatment-experienced (% of resistant) | resistant cases: naive (% of resistant) | TDR cases (% of resistant) | TDR cluster num. | TDR cluster sizes | ADR cases (% of resistant) | loss cases (% of resistant) |
|---|---|---|---|---|---|---|---|---|---|---|
| 14-03-16 | 39159 | A | 862 (2.2%) | 163 (18.9%) | 637 (73.9%) | 760.25 (88.2%) | 305.5 | 1-158 | 155.75 (18.1%) | 54 (6.3%) |
| 17-12-11 | 28011 | A | 582 (2.1%) | 124 (21.3%) | 428 (73.5%) | 485.75 (83.5%) | 226 | 1-89 | 122.25 (21.0%) | 26 (4.5%) |
| 17-12-06 | 13280 | A | 239 (1.8%) | 87 (36.4%) | 142 (59.4%) | 164.75 (68.9%) | 107.5 | 1-14 | 84.25 (35.3%) | 10 (4.2%) |
| 17-12-01 | 3195 | A | 61 (1.9%) | 31 (50.8%) | 29 (47.5%) | 33.50 (54.9%) | 28.5 | 1-4 | 29.50 (48.4%) | 2 (3.3%) |
| 14-11-96 | 7 | A | | | | | | | | |

**Table S9.** DRMs with prevalence > 0.5% found in position RT:E44 in B data set, and the evolution of their presence over time.

| date | total samples | DRM | resistant cases (% of all) | resistant cases: treatment-experienced (% of resistant) | resistant cases: naive (% of resistant) | TDR cases (% of resistant) | TDR cluster num. | TDR cluster sizes | ADR cases (% of resistant) | loss cases (% of resistant) |
|---|---|---|---|---|---|---|---|---|---|---|
| 14-03-16 | 39159 | D | 294 (0.8%) | 180 (61.2%) | 93 (31.6%) | 129.62 (44.1%) | 77 | 1-29 | 183.38 (62.4%) | 19 (6.5%) |
| 17-12-11 | 28011 | D | 249 (0.9%) | 170 (68.3%) | 60 (24.1%) | 87.50 (35.1%) | 58 | 1-9 | 176.50 (70.9%) | 15 (6.0%) |
| 17-12-06 | 13280 | D | 197 (1.5%) | 149 (75.6%) | 31 (15.7%) | 49.75 (25.3%) | 37.5 | 1-3 | 156.25 (79.3%) | 9 (4.6%) |
| 17-12-01 | 3195 | D | 79 (2.5%) | 66 (83.5%) | 7 (8.9%) | 11.00 (13.9%) | 10 | 1-2 | 70.00 (88.6%) | 2 (2.5%) |
| 14-11-96 | 7 | D | 1 (14.3%) | 1 (100.0%) | | | | | 1.00 (100.0%) | |

**Table S10.** DRMs with prevalence > 0.5% found in position RT:G190 in B data set, and the evolution of their presence over time.

| date | total samples | DRM | resistant cases (% of all) | resistant cases: treatment-experienced (% of resistant) | resistant cases: naive (% of resistant) | TDR cases (% of resistant) | TDR cluster num. | TDR cluster sizes | ADR cases (% of resistant) | loss cases (% of resistant) |
|---|---|---|---|---|---|---|---|---|---|---|
| 14-03-16 | 39159 | A | 447 (1.1%) | 342 (76.5%) | 68 (15.2%) | 117.25 (26.2%) | 97 | 1-6 | 350.75 (78.5%) | 21 (4.7%) |
| 17-12-11 | 28011 | A | 395 (1.4%) | 312 (79.0%) | 57 (14.4%) | 88.00 (22.3%) | 77 | 1-4 | 319.00 (80.8%) | 12 (3.0%) |
| 17-12-06 | 13280 | A | 341 (2.6%) | 280 (82.1%) | 38 (11.1%) | 60.00 (17.6%) | 58 | 1-2 | 287.00 (84.2%) | 6 (1.8%) |
| 17-12-01 | 3195 | A | 99 (3.1%) | 89 (89.9%) | 6 (6.1%) | 8.00 (8.1%) | 8 | 1-2 | 91.00 (91.9%) | |
| 14-11-96 | 7 | A | | | | | | | | |

**Table S11.** DRMs with prevalence > 0.5% found in position RT:H221 in B data set, and the evolution of their presence over time.

| date | total samples | DRM | resistant cases (% of all) | resistant cases: treatment-experienced (% of resistant) | resistant cases: treatment-naive (% of resistant) | TDR cases (% of resistant) | TDR cluster num. | TDR cluster sizes | ADR cases (% of resistant) | loss cases (% of resistant) |
|---|---|---|---|---|---|---|---|---|---|---|
| 14-03-16 | 39159 | Y | 475 (1.2%) | 269 (56.6%) | 162 (34.1%) | 220.00 (46.3%) | 87 | 1-64 | 267.00 (56.2%) | 12 (2.5%) |
| 17-12-11 | 28011 | Y | 403 (1.4%) | 240 (59.6%) | 132 (32.8%) | 169.50 (42.1%) | 65.5 | 1-49 | 242.50 (60.2%) | 9 (2.2%) |
| 17-12-06 | 13280 | Y | 251 (1.9%) | 185 (73.7%) | 48 (19.1%) | 66.50 (26.5%) | 43.5 | 1-9 | 188.50 (75.1%) | 4 (1.6%) |
| 17-12-01 | 3195 | Y | 61 (1.9%) | 47 (77.0%) | 11 (18.0%) | 12.50 (20.5%) | 12.5 | 1-1 | 48.50 (79.5%) | |
| 14-11-96 | 7 | Y | 1 (14.3%) | 1 (100.0%) | | | | | 1.00 (100.0%) | |

**Table S12.** DRMs with prevalence > 0.5% found in position RT:K101 in B data set, and the evolution of their presence over time.

| date | total samples | DRM | resistant cases (% of all) | resistant cases: treatment-experienced (% of resistant) | resistant cases: treatment-naive (% of resistant) | TDR cases (% of resistant) | TDR cluster num. | TDR cluster sizes | ADR cases (% of resistant) | loss cases (% of resistant) |
|---|---|---|---|---|---|---|---|---|---|---|
| 14-03-16 | 39159 | E | 276 (0.7%) | 189 (68.5%) | 64 (23.2%) | 94.50 (34.2%) | 71.5 | 1-9 | 190.50 (69.0%) | 9 (3.3%) |
| 17-12-11 | 28011 | E | 223 (0.8%) | 161 (72.2%) | 45 (20.2%) | 61.00 (27.4%) | 51.5 | 1-3 | 166.00 (74.4%) | 4 (1.8%) |
| 17-12-06 | 13280 | E | 176 (1.3%) | 141 (80.1%) | 24 (13.6%) | 35.50 (20.2%) | 31.5 | 1-2 | 143.50 (81.5%) | 3 (1.7%) |
| 17-12-01 | 3195 | E | 44 (1.4%) | 40 (90.9%) | 2 (4.5%) | 3.00 (6.8%) | 3 | 1-1 | 41.00 (93.2%) | |
| 14-11-96 | 7 | E | | | | | | | | |

**Table S13.** DRMs with prevalence > 0.5% found in position RT:K103 in B data set, and the evolution of their presence over time.

| date | total samples | DRM | resistant cases (% of all) | resistant cases: treatment-experienced (% of resistant) | resistant cases: treatment-naive (% of resistant) | TDR cases (% of resistant) | TDR cluster num. | TDR cluster sizes | ADR cases (% of resistant) | loss cases (% of resistant) |
|---|---|---|---|---|---|---|---|---|---|---|
| 14-03-16 | 39159 | N | 2025 (5.2%) | 1104 (54.5%) | 745 (36.8%) | 1071.51 (52.9%) | 516.5 | 1-78 | 1088.49 (53.8%) | 135 (6.7%) |
| 17-12-11 | 28011 | N | 1584 (5.7%) | 925 (58.4%) | 540 (34.1%) | 722.25 (45.6%) | 377.5 | 1-57 | 933.75 (58.9%) | 72 (4.5%) |
| 17-12-06 | 13280 | N | 1042 (7.8%) | 731 (70.2%) | 228 (21.9%) | 323.00 (31.0%) | 219 | 1-16 | 745.00 (71.5%) | 26 (2.5%) |
| 17-12-01 | 3195 | N | 269 (8.4%) | 230 (85.5%) | 24 (8.9%) | 33.50 (12.5%) | 33.5 | 1-2 | 235.50 (87.5%) | |
| 14-11-96 | 7 | N | | | | | | | | |

**Table S14.** DRMs with prevalence > 0.5% found in position RT:K219 in B data set, and the evolution of their presence over time.

| date | total samples | DRM | resistant cases (% of all) | resistant cases: treatment-experienced (% of resistant) | resistant cases: treatment-naive (% of resistant) | TDR cases (% of resistant) | TDR cluster num. | TDR cluster sizes | ADR cases (% of resistant) | loss cases (% of resistant) |
|---|---|---|---|---|---|---|---|---|---|---|
| | | E | 262 (0.7%) | 192 (73.3%) | 43 (16.4%) | 74.75 (28.5%) | 51.5 | 1-9 | 192.25 (73.4%) | 5 (1.9%) |
| 14-03-16 | 39159 | N | 238 (0.6%) | 92 (38.7%) | 127 (53.4%) | 161.00 (67.6%) | 23 | 1-113 | 81.00 (34.0%) | 4 (1.7%) |
| | | Q | 563 (1.4%) | 322 (57.2%) | 194 (34.5%) | 307.25 (54.6%) | 99 | 1-92 | 303.75 (54.0%) | 48 (8.5%) |
| | | E | 227 (0.8%) | 181 (79.7%) | 31 (13.7%) | 50.50 (22.2%) | 34 | 1-9 | 180.50 (79.5%) | 4 (1.8%) |
| 17-12-11 | 28011 | N | 194 (0.7%) | 83 (42.8%) | 101 (52.1%) | 119.50 (61.6%) | 17.5 | 1-86 | 76.50 (39.4%) | 2 (1.0%) |
| | | Q | 497 (1.8%) | 299 (60.2%) | 164 (33.0%) | 245.75 (49.4%) | 80.5 | 1-78 | 289.25 (58.2%) | 38 (7.6%) |
| | | E | 169 (1.3%) | 145 (85.8%) | 11 (6.5%) | 25.50 (15.1%) | 21.5 | 1-2 | 147.50 (87.3%) | 4 (2.4%) |
| 17-12-06 | 13280 | N | 101 (0.8%) | 66 (65.3%) | 32 (31.7%) | 40.50 (40.1%) | 13.5 | 1-17 | 62.50 (61.9%) | 2 (2.0%) |
| | | Q | 365 (2.7%) | 256 (70.1%) | 86 (23.6%) | 129.25 (35.4%) | 57 | 1-32 | 256.75 (70.3%) | 21 (5.8%) |
| | | E | 74 (2.3%) | 67 (90.5%) | 5 (6.8%) | 7.00 (9.5%) | 7 | 1-2 | 67.00 (90.5%) | |
| 17-12-01 | 3195 | N | 26 (0.8%) | 20 (76.9%) | 5 (19.2%) | 5.50 (21.2%) | 3.5 | 1-3 | 20.50 (78.8%) | |
| | | Q | 147 (4.6%) | 127 (86.4%) | 15 (10.2%) | 21.50 (14.6%) | 17.5 | 1-2 | 128.50 (87.4%) | 3 (2.0%) |
| | | E | | | | | | | | |
| 14-11-96 | 7 | N | 1 (14.3%) | 1 (100.0%) | | | | | 1.00 (100.0%) | |
| | | Q | 1 (14.3%) | 1 (100.0%) | | | | | 1.00 (100.0%) | |

**Table S15.** DRMs with prevalence > 0.5% found in position RT:K65 in B data set, and the evolution of their presence over time.

| date | total samples | DRM | resistant cases (% of all) | resistant cases: treatment-experienced (% of resistant) | resistant cases: treatment-naive (% of resistant) | TDR cases (% of resistant) | TDR cluster num. | TDR cluster sizes | ADR cases (% of resistant) | loss cases (% of resistant) |
|---|---|---|---|---|---|---|---|---|---|---|
| 14-03-16 | 39159 | R | 225 (0.6%) | 170 (75.6%) | 19 (8.4%) | 50.88 (22.6%) | 42 | 1-2 | 187.12 (83.2%) | 13 (5.8%) |
| 17-12-11 | 28011 | R | 189 (0.7%) | 146 (77.2%) | 15 (7.9%) | 37.62 (19.9%) | 33 | 1-2 | 159.38 (84.3%) | 8 (4.2%) |
| 17-12-06 | 13280 | R | 143 (1.1%) | 114 (79.7%) | 8 (5.6%) | 23.12 (16.2%) | 20.5 | 1-2 | 123.88 (86.6%) | 4 (2.8%) |
| 17-12-01 | 3195 | R | 19 (0.6%) | 18 (94.7%) | | 0.50 (2.6%) | 0.5 | 1-1 | 18.50 (97.4%) | |
| 14-11-96 | 7 | R | | | | | | | | |

**Table S16.** DRMs with prevalence > 0.5% found in position RT:K70 in B data set, and the evolution of their presence over time.

| date | total samples | DRM | resistant cases (% of all) | resistant cases: treatment-experienced (% of resistant) | resistant cases: treatment-naive (% of resistant) | TDR cases (% of resistant) | TDR cluster num. | TDR cluster sizes | ADR cases (% of resistant) | loss cases (% of resistant) |
|---|---|---|---|---|---|---|---|---|---|---|
| 14-03-16 | 39159 | R | 711 (1.8%) | 610 (85.8%) | 54 (7.6%) | 143.75 (20.2%) | 98.5 | 1-7 | 615.25 (86.5%) | 48 (6.8%) |
| 17-12-11 | 28011 | R | 681 (2.4%) | 596 (87.5%) | 46 (6.8%) | 120.50 (17.7%) | 84 | 1-5 | 602.50 (88.5%) | 42 (6.2%) |
| 17-12-06 | 13280 | R | 604 (4.5%) | 534 (88.4%) | 36 (6.0%) | 91.00 (15.1%) | 67 | 1-4 | 541.00 (89.6%) | 28 (4.6%) |
| 17-12-01 | 3195 | R | 294 (9.2%) | 269 (91.5%) | 18 (6.1%) | 27.50 (9.4%) | 25.5 | 1-2 | 269.50 (91.7%) | 3 (1.0%) |
| 14-11-96 | 7 | R | 2 (28.6%) | 2 (100.0%) | | | | | 2.00 (100.0%) | |

**Table S17.** DRMs with prevalence > 0.5% found in position RT:L210 in B data set, and the evolution of their presence over time.

| date | total samples | DRM | resistant cases (% of all) | treatment-experienced (% of resistant) | treatment-naive (% of resistant) | TDR cases (% of resistant) | TDR cluster num. | TDR cluster sizes | ADR cases (% of resistant) | loss cases (% of resistant) |
|---|---|---|---|---|---|---|---|---|---|---|
| 14-03-16 | 39159 | W | 705 (1.8%) | 520 (73.8%) | 140 (19.9%) | 205.00 (29.1%) | 147 | 1-9 | 524.00 (74.3%) | 24 (3.4%) |
| 17-12-11 | 28011 | W | 649 (2.3%) | 501 (77.2%) | 113 (17.4%) | 161.75 (24.9%) | 121 | 1-7 | 506.25 (78.0%) | 19 (2.9%) |
| 17-12-06 | 13280 | W | 556 (4.2%) | 452 (81.3%) | 70 (12.6%) | 106.75 (19.2%) | 81 | 1-7 | 460.25 (82.8%) | 11 (2.0%) |
| 17-12-01 | 3195 | W | 236 (7.4%) | 203 (86.0%) | 20 (8.5%) | 26.50 (11.2%) | 22.5 | 1-4 | 209.50 (88.8%) | |
| 14-11-96 | 7 | W | 3 (42.9%) | 3 (100.0%) | | | | | 3.00 (100.0%) | |

**Table S18.** DRMs with prevalence > 0.5% found in position RT:L74 in B data set, and the evolution of their presence over time.

| date | total samples | DRM | resistant cases (% of all) | treatment-experienced (% of resistant) | treatment-naive (% of resistant) | TDR cases (% of resistant) | TDR cluster num. | TDR cluster sizes | ADR cases (% of resistant) | loss cases (% of resistant) |
|---|---|---|---|---|---|---|---|---|---|---|
| 14-03-16 | 39159 | V | 242 (0.6%) | 200 (82.6%) | 17 (7.0%) | 38.25 (15.8%) | 36 | 1-3 | 207.75 (85.8%) | 4 (1.7%) |
| 17-12-11 | 28011 | V | 216 (0.8%) | 182 (84.3%) | 15 (6.9%) | 30.50 (14.1%) | 29.5 | 1-3 | 189.50 (87.7%) | 4 (1.9%) |
| 17-12-06 | 13280 | V | 172 (1.3%) | 147 (85.5%) | 11 (6.4%) | 21.50 (12.5%) | 21.5 | 1-2 | 152.50 (88.7%) | 2 (1.2%) |
| 17-12-01 | 3195 | V | 54 (1.7%) | 48 (88.9%) | 2 (3.7%) | 4.00 (7.4%) | 4 | 1-1 | 50.00 (92.6%) | |
| 14-11-96 | 7 | V | | | | | | | | |

**Table S19.** DRMs with prevalence > 0.5% found in position RT:M184 in B data set, and the evolution of their presence over time.

| date | total samples | DRM | resistant cases (% of all) | treatment-experienced (% of resistant) | treatment-naive (% of resistant) | TDR cases (% of resistant) | TDR cluster num. | TDR cluster sizes | ADR cases (% of resistant) | loss cases (% of resistant) |
|---|---|---|---|---|---|---|---|---|---|---|
| 14-03-16 | 39159 | V | 1899 (4.8%) | 1642 (86.5%) | 110 (5.8%) | 343.62 (18.1%) | 278.5 | 1-4 | 1667.38 (87.8%) | 112 (5.9%) |
| 17-12-11 | 28011 | V | 1703 (6.1%) | 1493 (87.7%) | 94 (5.5%) | 266.62 (15.7%) | 213.5 | 1-4 | 1517.38 (89.1%) | 81 (4.8%) |
| 17-12-06 | 13280 | V | 1428 (10.8%) | 1259 (88.2%) | 74 (5.2%) | 190.56 (13.3%) | 156.5 | 1-3 | 1284.44 (89.9%) | 47 (3.3%) |
| 17-12-01 | 3195 | V | 603 (18.9%) | 551 (91.4%) | 22 (3.6%) | 50.56 (8.4%) | 45 | 1-3 | 560.44 (92.9%) | 8 (1.3%) |
| 14-11-96 | 7 | V | 2 (28.6%) | 2 (100.0%) | | | | | 2.00 (100.0%) | |

**Table S20.** DRMs with prevalence > 0.5% found in position RT:M41 in B data set, and the evolution of their presence over time.

| date | total samples | DRM | resistant cases (% of all) | treatment-experienced (% of resistant) | treatment-naive (% of resistant) | TDR cases (% of resistant) | TDR cluster num. | TDR cluster sizes | ADR cases (% of resistant) | loss cases (% of resistant) |
|---|---|---|---|---|---|---|---|---|---|---|
| 14-03-16 | 39159 | L | 1513 (3.9%) | 982 (64.9%) | 428 (28.3%) | 618.50 (40.9%) | 305.5 | 1-55 | 968.50 (64.0%) | 74 (4.9%) |
| 17-12-11 | 28011 | L | 1389 (5.0%) | 938 (67.5%) | 367 (26.4%) | 507.50 (36.5%) | 266.5 | 1-38 | 935.50 (67.4%) | 54 (3.9%) |
| 17-12-06 | 13280 | L | 1099 (8.3%) | 826 (75.2%) | 208 (18.9%) | 294.00 (26.8%) | 188 | 1-16 | 830.00 (75.5%) | 25 (2.3%) |
| 17-12-01 | 3195 | L | 459 (14.4%) | 386 (84.1%) | 52 (11.3%) | 68.75 (15.0%) | 56 | 1-6 | 392.25 (85.5%) | 2 (0.4%) |
| 14-11-96 | 7 | L | 2 (28.6%) | 2 (100.0%) | | | | | 2.00 (100.0%) | |

**Table S21.** DRMs with prevalence > 0.5% found in position RT:S68 in B data set, and the evolution of their presence over time.

| date | total samples | DRM | resistant cases (% of all) | resistant cases: treatment-experienced (% of resistant) | resistant cases: treatment-naive (% of resistant) | TDR cases (% of resistant) | TDR cluster num. | TDR cluster sizes | ADR cases (% of resistant) | loss cases (% of resistant) |
|---|---|---|---|---|---|---|---|---|---|---|
| 14-03-16 | 39159 | G | 3178 (8.1%) | 436 (13.7%) | 2482 (78.1%) | 2922.56 (92.0%) | 612.5 | 1-759 | 316.44 (10.0%) | 61 (1.9%) |
| 17-12-11 | 28011 | G | 2055 (7.3%) | 318 (15.5%) | 1601 (77.9%) | 1845.00 (89.8%) | 468.5 | 1-486 | 251.00 (12.2%) | 41 (2.0%) |
| 17-12-06 | 13280 | G | 868 (6.5%) | 204 (23.5%) | 610 (70.3%) | 721.00 (83.1%) | 234.5 | 1-174 | 166.00 (19.1%) | 19 (2.2%) |
| 17-12-01 | 3195 | G | 148 (4.6%) | 63 (42.6%) | 78 (52.7%) | 98.50 (66.6%) | 60 | 1-22 | 54.50 (36.8%) | 5 (3.4%) |
| 14-11-96 | 7 | G | | | | | | | | |

**Table S22.** DRMs with prevalence > 0.5% found in position RT:T215 in B data set, and the evolution of their presence over time.

| date | total samples | DRM | resistant cases (% of all) | resistant cases: treatment-experienced (% of resistant) | resistant cases: treatment-naive (% of resistant) | TDR cases (% of resistant) | TDR cluster num. | TDR cluster sizes | ADR cases (% of resistant) | loss cases (% of resistant) |
|---|---|---|---|---|---|---|---|---|---|---|
| 14-03-16 | 39159 | D | 462 (1.2%) | 86 (18.6%) | 334 (72.3%) | 459.25 (99.4%) | 103 | 1-99 | 71.75 (15.5%) | 69 (14.9%) |
| | | F | 257 (0.7%) | 215 (83.7%) | 19 (7.4%) | 41.25 (16.1%) | 37 | 1-4 | 222.75 (86.7%) | 7 (2.7%) |
| | | S | 378 (1.0%) | 59 (15.6%) | 293 (77.5%) | 364.25 (96.4%) | 115 | 1-45 | 51.75 (13.7%) | 38 (10.1%) |
| | | Y | 883 (2.3%) | 790 (89.5%) | 37 (4.2%) | 119.50 (13.5%) | 102.5 | 1-5 | 785.50 (89.0%) | 22 (2.5%) |
| 17-12-11 | 28011 | D | 367 (1.3%) | 68 (18.5%) | 272 (74.1%) | 366.50 (99.9%) | 91 | 1-68 | 62.50 (17.0%) | 62 (16.9%) |
| | | F | 246 (0.9%) | 209 (85.0%) | 17 (6.9%) | 36.50 (14.8%) | 33.5 | 1-4 | 214.50 (87.2%) | 5 (2.0%) |
| | | S | 282 (1.0%) | 37 (13.1%) | 228 (80.9%) | 276.75 (98.1%) | 90 | 1-30 | 38.25 (13.6%) | 33 (11.7%) |
| | | Y | 858 (3.1%) | 770 (89.7%) | 36 (4.2%) | 105.00 (12.2%) | 93 | 1-5 | 768.00 (89.5%) | 15 (1.7%) |
| 17-12-06 | 13280 | D | 198 (1.5%) | 44 (22.2%) | 143 (72.2%) | 187.00 (94.4%) | 63 | 1-34 | 48.00 (24.2%) | 37 (18.7%) |
| | | F | 221 (1.7%) | 187 (84.6%) | 16 (7.2%) | 30.50 (13.8%) | 29.5 | 1-3 | 193.50 (87.6%) | 3 (1.4%) |
| | | S | 128 (1.0%) | 24 (18.8%) | 93 (72.7%) | 125.50 (98.0%) | 50.5 | 1-10 | 28.50 (22.3%) | 26 (20.3%) |
| | | Y | 777 (5.9%) | 696 (89.6%) | 33 (4.2%) | 82.00 (10.6%) | 73 | 1-5 | 701.00 (90.2%) | 6 (0.8%) |
| 17-12-01 | 3195 | D | 47 (1.5%) | 13 (27.7%) | 31 (66.0%) | 35.50 (75.5%) | 21.5 | 1-4 | 14.50 (30.9%) | 3 (6.4%) |
| | | F | 98 (3.1%) | 91 (92.9%) | 5 (5.1%) | 8.00 (8.2%) | 8 | 1-2 | 90.00 (91.8%) | |
| | | S | 23 (0.7%) | 7 (30.4%) | 14 (60.9%) | 17.00 (73.9%) | 9.5 | 1-3 | 9.00 (39.1%) | 3 (13.0%) |
| | | Y | 362 (11.3%) | 334 (92.3%) | 11 (3.0%) | 21.75 (6.0%) | 21.5 | 1-2 | 341.25 (94.3%) | 1 (0.3%) |
| 14-11-96 | 7 | D | 1 (14.3%) | 1 (100.0%) | | | | | 1.00 (100.0%) | |
| | | F | 1 (14.3%) | 1 (100.0%) | | | | | 1.00 (100.0%) | |
| | | S | | | | | | | | |
| | | Y | 2 (28.6%) | 2 (100.0%) | | | | | 2.00 (100.0%) | |

**Table S23.** DRMs with prevalence > 0.5% found in position RT:V106 in B data set, and the evolution of their presence over time.

| date | total samples | DRM | resistant cases (% of all) | resistant cases: treatment-experienced (% of resistant) | resistant cases: treatment-naive (% of resistant) | TDR cases (% of resistant) | TDR cluster num. | TDR cluster sizes | ADR cases (% of resistant) | loss cases (% of resistant) |
|---|---|---|---|---|---|---|---|---|---|---|
| 14-03-16 | 39159 | I | 1051 (2.7%) | 217 (20.6%) | 715 (68.0%) | 956.16 (91.0%) | 494.5 | 1-74 | 230.84 (22.0%) | 136 (12.9%) |
| 17-12-11 | 28011 | I | 717 (2.6%) | 152 (21.2%) | 494 (68.9%) | 651.38 (90.8%) | 340 | 1-57 | 164.62 (23.0%) | 99 (13.8%) |
| 17-12-06 | 13280 | I | 321 (2.4%) | 94 (29.3%) | 201 (62.6%) | 262.75 (81.9%) | 158 | 1-16 | 100.25 (31.2%) | 42 (13.1%) |
| 17-12-01 | 3195 | I | 64 (2.0%) | 29 (45.3%) | 35 (54.7%) | 45.00 (70.3%) | 31 | 1-3 | 29.00 (45.3%) | 10 (15.6%) |
| 14-11-96 | 7 | I | | | | | | | | |

**Table S24.** DRMs with prevalence > 0.5% found in position RT:V108 in B data set, and the evolution of their presence over time.

| date | total samples | DRM | resistant cases (% of all) | resistant cases treatment-experienced (% of resistant) | resistant cases treatment-naive (% of resistant) | TDR cases (% of resistant) | TDR cluster num. | TDR cluster sizes | ADR cases (% of resistant) | loss cases (% of resistant) |
|---|---|---|---|---|---|---|---|---|---|---|
| 14-03-16 | 39159 | I | 429 (1.1%) | 219 (51.0%) | 167 (38.9%) | 230.00 (53.6%) | 166 | 1-8 | 232.00 (54.1%) | 33 (7.7%) |
| 17-12-11 | 28011 | I | 338 (1.2%) | 186 (55.0%) | 126 (37.3%) | 161.25 (47.7%) | 127.5 | 1-4 | 195.75 (57.9%) | 19 (5.6%) |
| 17-12-06 | 13280 | I | 224 (1.7%) | 134 (59.8%) | 73 (32.6%) | 89.50 (40.0%) | 76.5 | 1-4 | 139.50 (62.3%) | 5 (2.2%) |
| 17-12-01 | 3195 | I | 53 (1.7%) | 36 (67.9%) | 13 (24.5%) | 15.00 (28.3%) | 14 | 1-2 | 38.00 (71.7%) | |
| 14-11-96 | 7 | I | | | | | | | | |

**Table S25.** DRMs with prevalence > 0.5% found in position RT:V179 in B data set, and the evolution of their presence over time.

| date | total samples | DRM | resistant cases (% of all) | resistant cases treatment-experienced (% of resistant) | resistant cases treatment-naive (% of resistant) | TDR cases (% of resistant) | TDR cluster num. | TDR cluster sizes | ADR cases (% of resistant) | loss cases (% of resistant) |
|---|---|---|---|---|---|---|---|---|---|---|
| 14-03-16 | 39159 | D | 790 (2.0%) | 151 (19.1%) | 559 (70.8%) | 694.12 (87.9%) | 334.5 | 1-45 | 158.88 (20.1%) | 63 (8.0%) |
| 17-12-11 | 28011 | D | 539 (1.9%) | 112 (20.8%) | 371 (68.8%) | 462.06 (85.7%) | 239 | 1-38 | 122.94 (22.8%) | 46 (8.5%) |
| 17-12-06 | 13280 | D | 225 (1.7%) | 76 (33.8%) | 128 (56.9%) | 165.50 (73.6%) | 105 | 1-23 | 82.50 (36.7%) | 23 (10.2%) |
| 17-12-01 | 3195 | D | 41 (1.3%) | 23 (56.1%) | 16 (39.0%) | 20.00 (48.8%) | 16 | 1-3 | 23.00 (56.1%) | 2 (4.9%) |
| 14-11-96 | 7 | D | | | | | | | | |

**Table S26.** DRMs with prevalence > 0.5% found in position RT:Y181 in B data set, and the evolution of their presence over time.

| date | total samples | DRM | resistant cases (% of all) | resistant cases treatment-experienced (% of resistant) | resistant cases treatment-naive (% of resistant) | TDR cases (% of resistant) | TDR cluster num. | TDR cluster sizes | ADR cases (% of resistant) | loss cases (% of resistant) |
|---|---|---|---|---|---|---|---|---|---|---|
| 14-03-16 | 39159 | C | 694 (1.8%) | 495 (71.3%) | 115 (16.6%) | 208.00 (30.0%) | 148 | 1-12 | 509.00 (73.3%) | 23 (3.3%) |
| 17-12-11 | 28011 | C | 600 (2.1%) | 442 (73.7%) | 96 (16.0%) | 159.50 (26.6%) | 110 | 1-12 | 457.50 (76.2%) | 17 (2.8%) |
| 17-12-06 | 13280 | C | 459 (3.5%) | 353 (76.9%) | 56 (12.2%) | 96.50 (21.0%) | 79.5 | 1-5 | 370.50 (80.7%) | 8 (1.7%) |
| 17-12-01 | 3195 | C | 126 (3.9%) | 112 (88.9%) | 7 (5.6%) | 10.50 (8.3%) | 10.5 | 1-1 | 115.50 (91.7%) | |
| 14-11-96 | 7 | C | | | | | | | | |

**Table S27.** DRMs with prevalence > 0.5% found in position PR:L90 in C data set, and the evolution of their presence over time.

| date | total samples | DRM | resistant cases (% of all) | resistant cases treatment-experienced (% of resistant) | resistant cases treatment-naive (% of resistant) | TDR cases (% of resistant) | TDR cluster num. | TDR cluster sizes | ADR cases (% of resistant) | loss cases (% of resistant) |
|---|---|---|---|---|---|---|---|---|---|---|
| 14-03-16 | 18809 | M | 108 (0.6%) | 62 (57.4%) | 22 (20.4%) | 43.00 (39.8%) | 35.5 | 1-4 | 67.00 (62.0%) | 2 (1.9%) |
| 17-12-11 | 14035 | M | 91 (0.6%) | 57 (62.6%) | 14 (15.4%) | 31.25 (34.3%) | 27 | 1-3 | 60.75 (66.8%) | 1 (1.1%) |
| 17-12-06 | 5374 | M | 68 (1.3%) | 47 (69.1%) | 6 (8.8%) | 16.00 (23.5%) | 15 | 1-3 | 52.00 (76.5%) | |
| 17-12-01 | 338 | M | 11 (3.3%) | 7 (63.6%) | 1 (9.1%) | 2.50 (22.7%) | 2.5 | 1-1 | 8.50 (77.3%) | |

**Table S28.** DRMs with prevalence > 0.5% found in position PR:Q58 in C data set, and the evolution of their presence over time.

| date | total samples | DRM | resistant cases (% of all) | resistant cases: treatment-experienced (% of resistant) | resistant cases: treatment-naive (% of resistant) | TDR cases (% of resistant) | TDR cluster num. | TDR cluster sizes | ADR cases (% of resistant) | loss cases (% of resistant) |
|---|---|---|---|---|---|---|---|---|---|---|
| 14-03-16 | 18809 | E | 153 (0.8%) | 31 (20.3%) | 97 (63.4%) | 117.75 (77.0%) | 81.5 | 1-12 | 37.25 (24.3%) | 2 (1.3%) |
| 17-12-11 | 14035 | E | 104 (0.7%) | 24 (23.1%) | 68 (65.4%) | 79.00 (76.0%) | 61 | 1-9 | 27.00 (26.0%) | 2 (1.9%) |
| 17-12-06 | 5374 | E | 34 (0.6%) | 10 (29.4%) | 20 (58.8%) | 23.00 (67.6%) | 21 | 1-2 | 12.00 (35.3%) | 1 (2.9%) |
| 17-12-01 | 338 | E | 1 (0.3%) | | | 0.50 (50.0%) | 0.5 | 1-1 | 0.50 (50.0%) | |

**Table S29.** DRMs with prevalence > 0.5% found in position RT:A98 in C data set, and the evolution of their presence over time.

| date | total samples | DRM | resistant cases (% of all) | resistant cases: treatment-experienced (% of resistant) | resistant cases: treatment-naive (% of resistant) | TDR cases (% of resistant) | TDR cluster num. | TDR cluster sizes | ADR cases (% of resistant) | loss cases (% of resistant) |
|---|---|---|---|---|---|---|---|---|---|---|
| 14-03-16 | 18809 | G | 239 (1.3%) | 115 (48.1%) | 78 (32.6%) | 112.12 (46.9%) | 99 | 1-4 | 126.88 (53.1%) | |
| 17-12-11 | 14035 | G | 187 (1.3%) | 89 (47.6%) | 65 (34.8%) | 88.12 (47.1%) | 77.5 | 1-4 | 98.88 (52.9%) | |
| 17-12-06 | 5374 | G | 93 (1.7%) | 55 (59.1%) | 17 (18.3%) | 32.31 (34.7%) | 28.5 | 1-4 | 60.69 (65.3%) | |
| 17-12-01 | 338 | G | 10 (3.0%) | 7 (70.0%) | 1 (10.0%) | 2.25 (22.5%) | 2 | 1-2 | 7.75 (77.5%) | |

**Table S30.** DRMs with prevalence > 0.5% found in position RT:D67 in C data set, and the evolution of their presence over time.

| date | total samples | DRM | resistant cases (% of all) | resistant cases: treatment-experienced (% of resistant) | resistant cases: treatment-naive (% of resistant) | TDR cases (% of resistant) | TDR cluster num. | TDR cluster sizes | ADR cases (% of resistant) | loss cases (% of resistant) |
|---|---|---|---|---|---|---|---|---|---|---|
| 14-03-16 | 18809 | N | 289 (1.5%) | 215 (74.4%) | 25 (8.7%) | 65.25 (22.6%) | 56.5 | 1-4 | 228.75 (79.2%) | 5 (1.7%) |
| 17-12-11 | 14035 | N | 247 (1.8%) | 187 (75.7%) | 22 (8.9%) | 49.00 (19.8%) | 42.5 | 1-4 | 199.00 (80.6%) | 1 (0.4%) |
| 17-12-06 | 5374 | N | 166 (3.1%) | 130 (78.3%) | 11 (6.6%) | 28.81 (17.4%) | 26.5 | 1-4 | 138.19 (83.2%) | 1 (0.6%) |
| 17-12-01 | 338 | N | 23 (6.8%) | 18 (78.3%) | 1 (4.3%) | 4.25 (18.5%) | 4 | 1-2 | 18.75 (81.5%) | |

**Table S31.** DRMs with prevalence > 0.5% found in position RT:E138 in C data set, and the evolution of their presence over time.

| date | total samples | DRM | resistant cases (% of all) | resistant cases: treatment-experienced (% of resistant) | resistant cases: treatment-naive (% of resistant) | TDR cases (% of resistant) | TDR cluster num. | TDR cluster sizes | ADR cases (% of resistant) | loss cases (% of resistant) |
|---|---|---|---|---|---|---|---|---|---|---|
| 14-03-16 | 18809 | A | 2176 (11.6%) | 512 (23.5%) | 1381 (63.5%) | 2136.88 (98.2%) | 415.5 | 1-1178 | 196.12 (9.0%) | 157 (7.2%) |
| 17-12-11 | 14035 | A | 1609 (11.5%) | 331 (20.6%) | 1132 (70.4%) | 1609.25 (100.0%) | 345 | 1-852 | 135.75 (8.4%) | 136 (8.5%) |
| 17-12-06 | 5374 | A | 617 (11.5%) | 154 (25.0%) | 417 (67.6%) | 615.50 (99.8%) | 150.5 | 1-322 | 65.50 (10.6%) | 64 (10.4%) |
| 17-12-01 | 338 | A | 28 (8.3%) | 10 (35.7%) | 12 (42.9%) | 26.50 (94.6%) | 5.5 | 1-18 | 6.50 (23.2%) | 5 (17.9%) |

**Table S32.** DRMs with prevalence > 0.5% found in position RT:G190 in C data set, and the evolution of their presence over time.

| date | total samples | DRM | resistant cases (% of all) | resistant cases treatment-experienced (% of resistant) | resistant cases naive (% of resistant) | TDR cases (% of resistant) | TDR cluster num. | TDR cluster sizes | ADR cases (% of resistant) | loss cases (% of resistant) |
|---|---|---|---|---|---|---|---|---|---|---|
| 14-03-16 | 18809 | A | 287 (1.5%) | 213 (74.2%) | 34 (11.8%) | 71.25 (24.8%) | 65.5 | 1-4 | 224.75 (78.3%) | 9 (3.1%) |
| 17-12-11 | 14035 | A | 233 (1.7%) | 173 (74.2%) | 27 (11.6%) | 50.75 (21.8%) | 47 | 1-4 | 184.25 (79.1%) | 2 (0.9%) |
| 17-12-06 | 5374 | A | 144 (2.7%) | 111 (77.1%) | 14 (9.7%) | 27.50 (19.1%) | 27 | 1-2 | 117.50 (81.6%) | 1 (0.7%) |
| 17-12-01 | 338 | A | 16 (4.7%) | 13 (81.2%) | 1 (6.2%) | 3.00 (18.8%) | 3 | 1-2 | 13.00 (81.2%) | |

**Table S33.** DRMs with prevalence > 0.5% found in position RT:H221 in C data set, and the evolution of their presence over time.

| date | total samples | DRM | resistant cases (% of all) | resistant cases treatment-experienced (% of resistant) | resistant cases naive (% of resistant) | TDR cases (% of resistant) | TDR cluster num. | TDR cluster sizes | ADR cases (% of resistant) | loss cases (% of resistant) |
|---|---|---|---|---|---|---|---|---|---|---|
| 14-03-16 | 18809 | Y | 173 (0.9%) | 123 (71.1%) | 27 (15.6%) | 45.75 (26.4%) | 42.5 | 1-2 | 133.25 (77.0%) | 6 (3.5%) |
| 17-12-11 | 14035 | Y | 144 (1.0%) | 102 (70.8%) | 23 (16.0%) | 36.25 (25.2%) | 34 | 1-2 | 111.75 (77.6%) | 4 (2.8%) |
| 17-12-06 | 5374 | Y | 79 (1.5%) | 57 (72.2%) | 14 (17.7%) | 19.75 (25.0%) | 19.5 | 1-2 | 60.25 (76.3%) | 1 (1.3%) |
| 17-12-01 | 338 | Y | 5 (1.5%) | 5 (100.0%) | | | | | 5.00 (100.0%) | |

**Table S34.** DRMs with prevalence > 0.5% found in position RT:K101 in C data set, and the evolution of their presence over time.

| date | total samples | DRM | resistant cases (% of all) | resistant cases treatment-experienced (% of resistant) | resistant cases naive (% of resistant) | TDR cases (% of resistant) | TDR cluster num. | TDR cluster sizes | ADR cases (% of resistant) | loss cases (% of resistant) |
|---|---|---|---|---|---|---|---|---|---|---|
| 14-03-16 | 18809 | E | 244 (1.3%) | 164 (67.2%) | 54 (22.1%) | 82.75 (33.9%) | 73.5 | 1-4 | 168.25 (69.0%) | 7 (2.9%) |
| 17-12-11 | 14035 | E | 189 (1.3%) | 129 (68.3%) | 43 (22.8%) | 59.75 (31.6%) | 54.5 | 1-4 | 133.25 (70.5%) | 4 (2.1%) |
| 17-12-06 | 5374 | E | 98 (1.8%) | 76 (77.6%) | 13 (13.3%) | 20.50 (20.9%) | 20.5 | 1-2 | 78.50 (80.1%) | 1 (1.0%) |
| 17-12-01 | 338 | E | 11 (3.3%) | 9 (81.8%) | | 2.00 (18.2%) | 2 | 1-2 | 9.00 (81.8%) | |

**Table S35.** DRMs with prevalence > 0.5% found in position RT:K103 in C data set, and the evolution of their presence over time.

| date | total samples | DRM | resistant cases (% of all) | resistant cases treatment-experienced (% of resistant) | resistant cases naive (% of resistant) | TDR cases (% of resistant) | TDR cluster num. | TDR cluster sizes | ADR cases (% of resistant) | loss cases (% of resistant) |
|---|---|---|---|---|---|---|---|---|---|---|
| 14-03-16 | 18809 | N | 882 (4.7%) | 605 (68.6%) | 182 (20.6%) | 317.12 (36.0%) | 267 | 1-5 | 615.88 (69.8%) | 51 (5.8%) |
| 17-12-11 | 14035 | N | 654 (4.7%) | 449 (68.7%) | 139 (21.3%) | 204.62 (31.3%) | 184.5 | 1-3 | 472.38 (72.2%) | 23 (3.5%) |
| 17-12-06 | 5374 | N | 376 (7.0%) | 286 (76.1%) | 53 (14.1%) | 83.00 (22.1%) | 78 | 1-3 | 298.00 (79.3%) | 5 (1.3%) |
| 17-12-01 | 338 | N | 23 (6.8%) | 18 (78.3%) | 4 (17.4%) | 4.50 (19.6%) | 4.5 | 1-1 | 18.50 (80.4%) | |

**Table S36.** DRMs with prevalence > 0.5% found in position RT:K219 in C data set, and the evolution of their presence over time.

| date | total | DRM | resistant cases | | | TDR | | | ADR | loss |
|---|---|---|---|---|---|---|---|---|---|---|
| | samples | | (% of all) | treatment- | | cases | cluster | | cases | cases |
| | | | | experienced | naive | (% of | num. | sizes | (% of | (% of |
| | | | | (% of resistant) | | resistant) | | | resistant) | resistant) |
| 14-03-16 | 18809 | E | 109 (0.6%) | 80 (73.4%) | 15 (13.8%) | 25.50 (23.4%) | 24.5 | 1-3 | 83.50 (76.6%) | |
| 17-12-11 | 14035 | E | 85 (0.6%) | 65 (76.5%) | 12 (14.1%) | 17.00 (20.0%) | 17 | 1-2 | 68.00 (80.0%) | |
| 17-12-06 | 5374 | E | 54 (1.0%) | 42 (77.8%) | 6 (11.1%) | 10.00 (18.5%) | 10 | 1-2 | 44.00 (81.5%) | |
| 17-12-01 | 338 | E | 5 (1.5%) | 5 (100.0%) | | | | | 5.00 (100.0%) | |

**Table S37.** DRMs with prevalence > 0.5% found in position RT:K65 in C data set, and the evolution of their presence over time.

| date | total | DRM | resistant cases | | | TDR | | | ADR | loss |
|---|---|---|---|---|---|---|---|---|---|---|
| | samples | | (% of all) | treatment- | | cases | cluster | | cases | cases |
| | | | | experienced | naive | (% of | num. | sizes | (% of | (% of |
| | | | | (% of resistant) | | resistant) | | | resistant) | resistant) |
| 14-03-16 | 18809 | R | 244 (1.3%) | 199 (81.6%) | 15 (6.1%) | 38.50 (15.8%) | 36.5 | 1-2 | 211.50 (86.7%) | 6 (2.5%) |
| 17-12-11 | 14035 | R | 177 (1.3%) | 142 (80.2%) | 11 (6.2%) | 26.50 (15.0%) | 25.5 | 1-2 | 153.50 (86.7%) | 3 (1.7%) |
| 17-12-06 | 5374 | R | 97 (1.8%) | 79 (81.4%) | 3 (3.1%) | 13.00 (13.4%) | 13 | 1-2 | 86.00 (88.7%) | 2 (2.1%) |
| 17-12-01 | 338 | R | 1 (0.3%) | | | 0.50 (50.0%) | 0.5 | 1-1 | 0.50 (50.0%) | |

**Table S38.** DRMs with prevalence > 0.5% found in position RT:K70 in C data set, and the evolution of their presence over time.

| date | total | DRM | resistant cases | | | TDR | | | ADR | loss |
|---|---|---|---|---|---|---|---|---|---|---|
| | samples | | (% of all) | treatment- | | cases | cluster | | cases | cases |
| | | | | experienced | naive | (% of | num. | sizes | (% of | (% of |
| | | | | (% of resistant) | | resistant) | | | resistant) | resistant) |
| 14-03-16 | 18809 | R | 196 (1.0%) | 152 (77.6%) | 19 (9.7%) | 38.62 (19.7%) | 35.5 | 1-4 | 161.38 (82.3%) | 4 (2.0%) |
| 17-12-11 | 14035 | R | 175 (1.2%) | 140 (80.0%) | 16 (9.1%) | 28.62 (16.4%) | 27 | 1-2 | 149.38 (85.4%) | 3 (1.7%) |
| 17-12-06 | 5374 | R | 123 (2.3%) | 98 (79.7%) | 10 (8.1%) | 19.00 (15.4%) | 19 | 1-2 | 105.00 (85.4%) | 1 (0.8%) |
| 17-12-01 | 338 | R | 14 (4.1%) | 11 (78.6%) | 1 (7.1%) | 2.00 (14.3%) | 2 | 1-1 | 12.00 (85.7%) | |

**Table S39.** DRMs with prevalence > 0.5% found in position RT:M184 in C data set, and the evolution of their presence over time.

| date | total | DRM | resistant cases | | | TDR | | | ADR | loss |
|---|---|---|---|---|---|---|---|---|---|---|
| | samples | | (% of all) | treatment- | | cases | cluster | | cases | cases |
| | | | | experienced | naive | (% of | num. | sizes | (% of | (% of |
| | | | | (% of resistant) | | resistant) | | | resistant) | resistant) |
| 14-03-16 | 18809 | V | 1009 (5.4%) | 789 (78.2%) | 79 (7.8%) | 213.88 (21.2%) | 197 | 1-4 | 833.12 (82.6%) | 38 (3.8%) |
| 17-12-11 | 14035 | V | 817 (5.8%) | 642 (78.6%) | 70 (8.6%) | 154.38 (18.9%) | 144.5 | 1-4 | 680.62 (83.3%) | 18 (2.2%) |
| 17-12-06 | 5374 | V | 524 (9.8%) | 419 (80.0%) | 41 (7.8%) | 86.06 (16.4%) | 80 | 1-4 | 443.94 (84.7%) | 6 (1.1%) |
| 17-12-01 | 338 | V | 53 (15.7%) | 36 (67.9%) | 3 (5.7%) | 12.19 (23.0%) | 9 | 1-4 | 41.81 (78.9%) | 1 (1.9%) |

**Table S40.** DRMs with prevalence > 0.5% found in position RT:M41 in C data set, and the evolution of their presence over time.

| date | total samples | DRM | resistant cases (% of all) | resistant cases: treatment-experienced (% of resistant) | resistant cases: naive (% of resistant) | TDR cases (% of resistant) | TDR cluster num. | TDR cluster sizes | ADR cases (% of resistant) | loss cases (% of resistant) |
|---|---|---|---|---|---|---|---|---|---|---|
| 14-03-16 | 18809 | L | 171 (0.9%) | 117 (68.4%) | 25 (14.6%) | 51.72 (30.2%) | 43.5 | 1-5 | 120.28 (70.3%) | 1 (0.6%) |
| 17-12-11 | 14035 | L | 146 (1.0%) | 101 (69.2%) | 20 (13.7%) | 40.72 (27.9%) | 33.5 | 1-5 | 106.28 (72.8%) | 1 (0.7%) |
| 17-12-06 | 5374 | L | 106 (2.0%) | 79 (74.5%) | 9 (8.5%) | 22.28 (21.0%) | 19 | 1-5 | 83.72 (79.0%) | |
| 17-12-01 | 338 | L | 13 (3.8%) | 10 (76.9%) | | 2.75 (21.2%) | 2.5 | 1-2 | 10.25 (78.8%) | |

**Table S41.** DRMs with prevalence > 0.5% found in position RT:S68 in C data set, and the evolution of their presence over time.

| date | total samples | DRM | resistant cases (% of all) | resistant cases: treatment-experienced (% of resistant) | resistant cases: naive (% of resistant) | TDR cases (% of resistant) | TDR cluster num. | TDR cluster sizes | ADR cases (% of resistant) | loss cases (% of resistant) |
|---|---|---|---|---|---|---|---|---|---|---|
| 14-03-16 | 18809 | G | 160 (0.9%) | 51 (31.9%) | 87 (54.4%) | 103.25 (64.5%) | 77 | 1-12 | 57.75 (36.1%) | 1 (0.6%) |
| 17-12-11 | 14035 | G | 113 (0.8%) | 36 (31.9%) | 59 (52.2%) | 71.00 (62.8%) | 56.5 | 1-10 | 43.00 (38.1%) | 1 (0.9%) |
| 17-12-06 | 5374 | G | 37 (0.7%) | 16 (43.2%) | 15 (40.5%) | 19.00 (51.4%) | 15.5 | 1-4 | 19.00 (51.4%) | 1 (2.7%) |
| 17-12-01 | 338 | G | 1 (0.3%) | | 1 (100.0%) | 1.00 (100.0%) | 1 | 1-1 | | |

**Table S42.** DRMs with prevalence > 0.5% found in position RT:T215 in C data set, and the evolution of their presence over time.

| date | total samples | DRM | resistant cases (% of all) | resistant cases: treatment-experienced (% of resistant) | resistant cases: naive (% of resistant) | TDR cases (% of resistant) | TDR cluster num. | TDR cluster sizes | ADR cases (% of resistant) | loss cases (% of resistant) |
|---|---|---|---|---|---|---|---|---|---|---|
| 14-03-16 | 18809 | Y | 137 (0.7%) | 97 (70.8%) | 13 (9.5%) | 37.97 (27.7%) | 31.5 | 1-5 | 105.03 (76.7%) | 6 (4.4%) |
| 17-12-11 | 14035 | Y | 125 (0.9%) | 89 (71.2%) | 12 (9.6%) | 32.97 (26.4%) | 26 | 1-5 | 96.03 (76.8%) | 4 (3.2%) |
| 17-12-06 | 5374 | Y | 103 (1.9%) | 76 (73.8%) | 7 (6.8%) | 20.78 (20.2%) | 17.5 | 1-5 | 83.22 (80.8%) | 1 (1.0%) |
| 17-12-01 | 338 | Y | 17 (5.0%) | 14 (82.4%) | | 2.75 (16.2%) | 2.5 | 1-2 | 14.25 (83.8%) | |

**Table S43.** DRMs with prevalence > 0.5% found in position RT:V106 in C data set, and the evolution of their presence over time.

| date | total samples | DRM | resistant cases (% of all) | resistant cases: treatment-experienced (% of resistant) | resistant cases: naive (% of resistant) | TDR cases (% of resistant) | TDR cluster num. | TDR cluster sizes | ADR cases (% of resistant) | loss cases (% of resistant) |
|---|---|---|---|---|---|---|---|---|---|---|
| 14-03-16 | 18809 | M | 381 (2.0%) | 301 (79.0%) | 36 (9.4%) | 71.25 (18.7%) | 66 | 1-4 | 319.75 (83.9%) | 10 (2.6%) |
| 17-12-11 | 14035 | M | 285 (2.0%) | 225 (78.9%) | 31 (10.9%) | 47.25 (16.6%) | 46 | 1-3 | 238.75 (83.8%) | 1 (0.4%) |
| 17-12-06 | 5374 | M | 151 (2.8%) | 122 (80.8%) | 11 (7.3%) | 21.50 (14.2%) | 21.5 | 1-2 | 130.50 (86.4%) | 1 (0.7%) |
| 17-12-01 | 338 | M | 8 (2.4%) | 7 (87.5%) | | 1.50 (18.8%) | 1.5 | 1-2 | 6.50 (81.2%) | |

**Table S44.** DRMs with prevalence > 0.5% found in position RT:V108 in C data set, and the evolution of their presence over time.

| date | total samples | DRM | resistant cases (% of all) | treatment-experienced (% of resistant) | treatment-naive (% of resistant) | TDR cases (% of resistant) | TDR cluster num. | TDR cluster sizes | ADR cases (% of resistant) | loss cases (% of resistant) |
|---|---|---|---|---|---|---|---|---|---|---|
| 14-03-16 | 18809 | I | 194 (1.0%) | 114 (58.8%) | 55 (28.4%) | 77.75 (40.1%) | 72 | 1-3 | 123.25 (63.5%) | 7 (3.6%) |
| 17-12-11 | 14035 | I | 142 (1.0%) | 83 (58.5%) | 39 (27.5%) | 53.25 (37.5%) | 51 | 1-2 | 91.75 (64.6%) | 3 (2.1%) |
| 17-12-06 | 5374 | I | 70 (1.3%) | 47 (67.1%) | 13 (18.6%) | 20.00 (28.6%) | 19 | 1-2 | 51.00 (72.9%) | 1 (1.4%) |
| 17-12-01 | 338 | I | 3 (0.9%) | 3 (100.0%) | | | | | 3.00 (100.0%) | |

**Table S45.** DRMs with prevalence > 0.5% found in position RT:V179 in C data set, and the evolution of their presence over time.

| date | total samples | DRM | resistant cases (% of all) | treatment-experienced (% of resistant) | treatment-naive (% of resistant) | TDR cases (% of resistant) | TDR cluster num. | TDR cluster sizes | ADR cases (% of resistant) | loss cases (% of resistant) |
|---|---|---|---|---|---|---|---|---|---|---|
| 14-03-16 | 18809 | D | 294 (1.6%) | 99 (33.7%) | 159 (54.1%) | 214.50 (73.0%) | 139 | 1-19 | 102.50 (34.9%) | 23 (7.8%) |
| | | E | 120 (0.6%) | 11 (9.2%) | 34 (28.3%) | 108.25 (90.2%) | 24 | 1-80 | 12.75 (10.6%) | 1 (0.8%) |
| 17-12-11 | 14035 | D | 213 (1.5%) | 71 (33.3%) | 119 (55.9%) | 146.50 (68.8%) | 101 | 1-10 | 78.50 (36.9%) | 12 (5.6%) |
| | | E | 37 (0.3%) | 5 (13.5%) | 19 (51.4%) | 30.25 (81.8%) | 15 | 1-15 | 6.75 (18.2%) | |
| 17-12-06 | 5374 | D | 86 (1.6%) | 32 (37.2%) | 42 (48.8%) | 50.75 (59.0%) | 39.5 | 1-5 | 36.25 (42.2%) | 1 (1.2%) |
| | | E | 8 (0.1%) | 4 (50.0%) | 3 (37.5%) | 3.50 (43.8%) | 3.5 | 1-1 | 4.50 (56.2%) | |
| 17-12-01 | 338 | D | 10 (3.0%) | 4 (40.0%) | 3 (30.0%) | 5.50 (55.0%) | 5.5 | 1-2 | 4.50 (45.0%) | |
| | | E | | | | | | | | |

**Table S46.** DRMs with prevalence > 0.5% found in position RT:Y181 in C data set, and the evolution of their presence over time.

| date | total samples | DRM | resistant cases (% of all) | treatment-experienced (% of resistant) | treatment-naive (% of resistant) | TDR cases (% of resistant) | TDR cluster num. | TDR cluster sizes | ADR cases (% of resistant) | loss cases (% of resistant) |
|---|---|---|---|---|---|---|---|---|---|---|
| 14-03-16 | 18809 | C | 419 (2.2%) | 299 (71.4%) | 56 (13.4%) | 108.38 (25.9%) | 98 | 1-4 | 321.62 (76.8%) | 11 (2.6%) |
| 17-12-11 | 14035 | C | 334 (2.4%) | 234 (70.1%) | 50 (15.0%) | 80.62 (24.1%) | 76.5 | 1-3 | 256.38 (76.8%) | 3 (0.9%) |
| 17-12-06 | 5374 | C | 183 (3.4%) | 133 (72.7%) | 21 (11.5%) | 38.00 (20.8%) | 36 | 1-2 | 146.00 (79.8%) | 1 (0.5%) |
| 17-12-01 | 338 | C | 17 (5.0%) | 13 (76.5%) | | 2.00 (11.8%) | 2 | 1-1 | 15.00 (88.2%) | |
